# Dual-wavelength time-of-flight interferometric speckle-contrast optical spectroscopy (dual-wavelength TOF-iSCOS) for co-registered blood-flow and hemoglobin sensing

**DOI:** 10.64898/2026.09.02.26362024

**Authors:** Klaudia Nowacka-Pieszak, Marcin Marzejon, Neda Mogharari, Dawid Borycki

**Affiliations:** International Centre for Translational Eye Research, Skierniewicka 10A, 01-230 Warsaw, Poland; Institute of Physical Chemistry, Polish Academy of Sciences, M. Kasprzaka 44/52, 01-224 Warsaw, Poland

**Keywords:** dual-wavelength interferometric field correlation, time-of-flight, wavelength multiplexing, first-order autocorrelation, finite-integration interferometric visibility, decorrelation rate, blood-flow index, modified Beer-Lambert law

## Abstract

Diffuse optical measurements of blood flow, blood volume, and blood oxygenation often use separate devices and photon populations. Combining these contrasts within a common photon population would enable their direct co-registration while retaining time-of-flight-based depth discrimination. We introduce dual-wavelength time-of-flight interferometric speckle-contrast optical spectroscopy (TOF-iSCOS), built on an interferometric near-infrared spectroscopy (iNIRS) architecture that temporally multiplexes 780 and 852 nm swept lasers through a shared detection channel. It recovers co-registered flow-sensitive field dynamics and wavelength-resolved attenuation from the same complex-field acquisition. Three 50 ps TOF gates sample increasingly late-arriving photons. Within each gate, *g*_1_ was calculated directly as the normalized temporal autocorrelation of the reconstructed complex field. *κ*^2^*(T)* was reconstructed by finite-time integration of measured *g*_1_ and finite-integration fits yielded decorrelation rates. No camera speckle-variance contrast or Siegert substitution was used. Gate-integrated TPSF intensities yielded optical-density changes *ΔOD*, and division by the baseline-TPSF-weighted mean photon path length gave effective *Δµ*_*a*_ for two-wavelength hemoglobin inversion. In 12 forearm cuff-occlusion recordings from 11 adults, TOF-iSCOS detected flow suppression during occlusion and reactive hyperemia after release at both wavelengths. At 780 nm, the Late-minus-Early reactive-hyperemia amplitude in the relative blood-flow index (rBFI) was +0.647 (95% CI 0.194 to 1.124), whereas the Early-minus-Late cuff *Δµ*_*a*_ contrast was +0.196 cm^−1^ (95% CI 0.123 to 0.280). In the photon-limited 852 nm Late gate, the accepted-frame fraction was 0.994 for *κ*^2^-based fitting versus 0.831 for *g*_1_-based fitting. Exploratory phase-resolved analysis suggested that the initial hemoglobin response was concentrated in early-arriving photons, while later responses differed in magnitude and direction. Wavelength, field dynamics, and photon time-of-flight thus provide complementary physiological information; the largest response need not identify the phase of interest.

## 1. Introduction

Blood flow, blood volume and blood oxygenation are coupled but non-interchangeable indicators of tissue function. Their joint evolution across time and depth, including how perfusion, volume and oxygen extraction change together, can be more informative than any one quantity alone. Diffuse optical methods recover these quantities through two principal measurement contrasts. Temporal fluctuations in multiply scattered coherent light reflect red-blood-cell motion, and diffuse correlation spectroscopy (DCS) converts the resulting field decorrelation into a microvascular blood-flow index [1]. DCS is now an established approach for cerebral and peripheral perfusion monitoring [2]. Spectral attenuation, by contrast, reports changes in hemoglobin concentration. Near-infrared spectroscopy commonly estimates these changes using the modified Beer–Lambert law [3], with a differential path-length factor accounting for the wavelength- and tissue-dependent photon path length [4], within the broader framework of diffuse optical monitoring and tomography [5]. Although these contrasts are complementary, they are generally acquired using separate instruments or, when co-located, through different source-detector geometries, photon populations or acquisition clocks. Their coupling is therefore inferred across measurements rather than observed within the same photon population.

Photon time-of-flight provides an additional encoding dimension related to photon path length and depth sensitivity. Time-resolved reflectance first established that the distribution of photon arrival times encodes tissue optical properties [6]. Time-domain diffuse optics has since developed into a family of depth-sensitive techniques [7], including time-domain functional NIRS for cortical hemoglobin mapping [8]. The same principle has also been applied to blood-flow measurements. Time-domain DCS resolves correlation decay by photon arrival time [9], enabling depth-resolved blood-flow measurements *in vivo* [10], although the recovered decorrelation depends on the instrument response and temporal-gate width [11]. Interferometric detection further extended this approach, with time-of-flight-resolved light-field fluctuations revealing deep-tissue physiology [12]. Monte Carlo analysis also indicates that later-arriving photons can provide greater selectivity for pulsatile cerebral flow [13]. These approaches, however, generally apply time-of-flight resolution to either hemodynamic attenuation or flow contrast, rather than recovering both from the same detected field.

A parallel approach estimates blood flow from the visibility of a finite-exposure speckle pattern rather than from its full temporal autocorrelation [14,15]. Speckle-contrast optical spectroscopy (SCOS) extended this principle to diffuse, deep-tissue measurements [16] and was subsequently adapted for time-gated measurements at quasi-null source-detector separation [17] and benchmarked against DCS [18]. Interferometric, or heterodyne, variants have further improved sensitivity and photon efficiency for cerebral monitoring and portable instrumentation [19–21]. Most implementations, however, remain focused on flow and operate at a single wavelength.

Interferometric near-infrared spectroscopy (iNIRS) provides a promising basis for a unified measurement because it records the complex optical field rather than intensity alone. Swept-source heterodyne detection resolves this field by wavelength and photon time of flight, allowing both optical and dynamical tissue properties to be estimated [22]. Direct field detection also enables calculation of the first-order field autocorrelation, *g*_1_, without inferring it from an intensity autocorrelation through the Siegert relation [23]. Reflectance-mode iNIRS has recovered absorption, scattering and a blood-flow index in the mouse brain in vivo [24]. Its flow estimates also depend on wavelength [25]. Rather than treating this dependence solely as a confound, wavelength-multiplexed acquisition can preserve channel-specific flow information while using dual-wavelength spectral attenuation to recover hemoglobin contrast.

Recent work has extended interferometric speckle-contrast measurements to the time-of-flight domain. Time-of-flight-resolved interferometric speckle-contrast optical spectroscopy (TOF-iSCOS) was first introduced in a conference report [26] and subsequently developed for depth-resolved flow sensing [27]. A field-level finite-integration interferometric visibility formulation, was formalized in continuous-wave parallel iNIRS [28], and a hybrid iNIRS implementation later demonstrated time-of-flight-resolved flow monitoring in humans [29]. Together, these studies established the basis for interferometric, time-of-flight-resolved flow sensing. What remains missing is a single acquisition that co-registers spectral attenuation, field dynamics and photon time of flight, thereby combining hemoglobin, flow and path-length information within the same detected field.

Achieving this measurement requires interleaving the wavelengths while preserving wavelength-specific sweep linearization, time-of-flight reconstruction, gate definition and temporal correlation sampling. Here, we introduce dual-wavelength TOF-iSCOS, a temporally multiplexed interferometric modality implemented with swept sources at 780 and 852 nm with a shared probe, detector and acquisition clock (Fig. 1). The multiplexed record is separated by wavelength, and each complex field is independently reconstructed along the time-of-flight axis. Flow dynamics are estimated using both the measured field autocorrelation and reconstructed finite-integration interferometric visibility, yielding-based and -based decorrelation-rate estimates without requiring a Siegert conversion. Two-wavelength gate intensities provide path-length-normalized attenuation and a secondary hemoglobin inversion. The same wavelength-multiplexed architecture is extendable to additional wavelengths, but the present validation is explicitly dual-wavelength.

**Figure 1.**
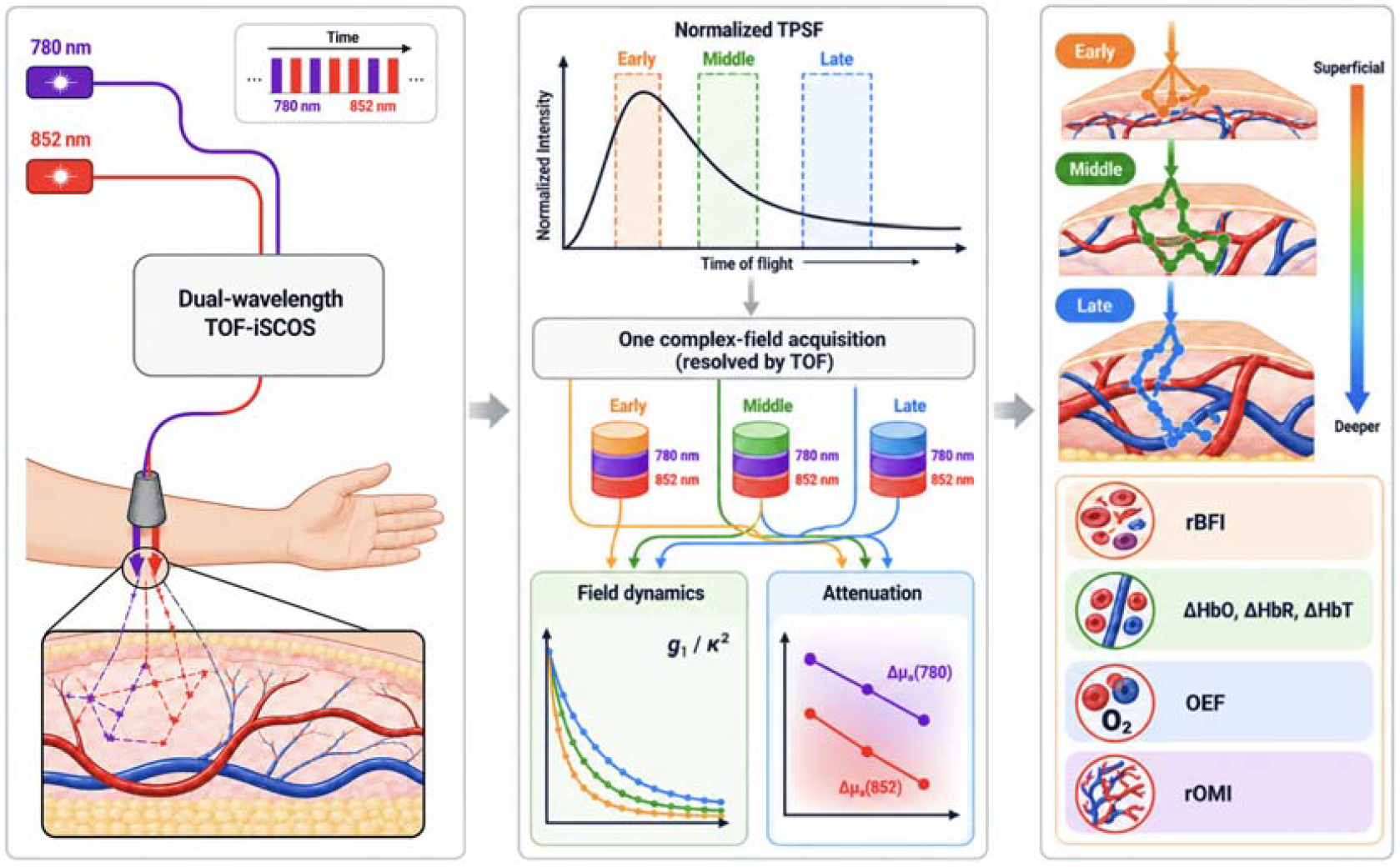
Conceptual overview of dual-wavelength TOF-iSCOS. Temporally multiplexed 780- and 852 nm illumination is measured using a shared interferometric system and a single detection channel. The reconstructed TPSF is sampled using fixed Early, Middle and Late time-of-flight gates, representing progressively later photon populations. Within each gate, field dynamics are quantified using direct- and reconstructed-observables, while wavelength-resolved attenuation provides at 780 and 852 nm. Their combination yields co-registered relative blood-flow index (rBFI), effective changes in oxyhemoglobin, deoxyhemoglobin and total hemoglobin (, and), oxygen-extraction fraction (OEF), and relative oxygen metabolic index (rOMI).

This methodological study establishes the instrument and wavelength-specific reconstruction framework and validates both measurement branches using controlled forearm cuff occlusion in vivo. The validation tests whether flow and spectroscopic responses remain physiologically responsive, stable and quantitatively interpretable when recovered from the same temporally multiplexed complex-field acquisition.

## 2. Theory

The forward model follows the interferometric near-infrared spectroscopy framework [22,23] and extends the single-wavelength TOF-iSCOS derivation [26–28] by treating wavelength explicitly. Here, *λ* ∈ {780,852 nmm} labels the nominal wavelength channel, *ν* denotes optical frequency within a sweep, *τ*_*s*_ is the photon time-of-flight, *τ*_*d*_ is the correlation lag, *t* denotes the slow monitoring time associated with each measurement, and *T* is the effective integration time. The detector provides a single balanced interferometric output. The operative dynamic observable is therefore the temporal correlation of the recovered optical field. Unlike camera-based speckle measurements, the present approach does not rely on a spatial ensemble of detector pixels or speckles.

### 2.1 Swept-source interferometric signal and the TOF-resolved optical field

For each wavelength *λ*, the source is swept over an optical-frequency bandwidth *Δν* while a detector records the spectral interferogram

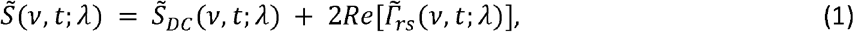

where 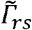 is the mutual coherence between the reference and sample fields, and 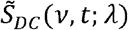 contains the reference and sample self-intensity terms. The spectral samples are linearized onto a uniform optical-frequency grid, equivalently a uniform vacuum-wavenumber grid *k*_*o*_ *=* 2*π ν*/*c*_*o*_ (*c*_*o*_ denoting the speed of light). An inverse Fourier transform along *ν* then yields the complex delay-resolved heterodyne field:

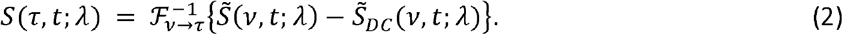

Only the positive-delay branch is retained, thereby excluding its conjugate mirror.

The raw Fourier-delay coordinate *τ* is calibrated independently for each wavelength. The peak position of the measured instrument response (Section 3.3) defines the zero-delay offset *τ*_*o,γ*_, and the calibrated photon time of flight is *τ*_*s=*_ *τ*− *τ*_*o,γ*_.

After this calibration, the recovered field is expressed as *S*(*τ*_*s*_,*t*;*λ*). Because the two swept sources may have different usable optical bandwidths, their time-of-flight resolutions may also differ, approximately following 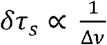 [29]. The resulting quantity *S*(*τ*_*s*_,*t*;*λ*) is the fundamental complex-field observable of the platform.

### 2.2 Field autocorrelation and the photon time-of-flight distribution

Repeated short-exposure sweeps provide the TOF-resolved first-order field autocorrelation. For each slow monitoring time *t*, the correlation is estimated over a local ensemble of sweeps

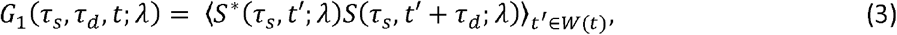

where *W*(*t*) denotes the local ensemble of sweeps associated with the measurement at monitoring time *t*.

The corresponding normalized field autocorrelation and zero-lag intensity are:

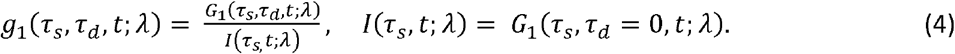

The zero-lag quantity *I*(*τ*_*s*_;*λ*) represents the wavelength-resolved photon time-of-flight distribution (DTOF), also referred to here as the temporal point-spread function (TPSF). Thus, the same interferometric measurement provides both the photon-path-length distribution and the TOF-resolved field dynamics.

For compactness, let Δ*τ*_*s*_ = [*τ*_*s,1*_, τ_*s,2*_] denote a finite TOF gate. The corresponding gate-resolved field autocorrelation is

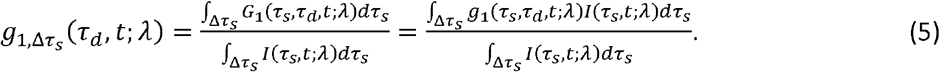

Thus, the gate-resolved autocorrelation is a TPSF-weighted average of the TOF-resolved field correlations. Because each finite gate contains a distribution of photon path lengths, its autocorrelation decay is generally a weighted mixture rather than an exact single exponential. Any fitted decay rate should therefore be interpreted as an effective gate-level parameter.

### 2.3 Finite-integration interferometric visibility and its relation to *g*_*1*_

To relate the recovered field autocorrelation to finite-integration interferometric visibility, we extend our earlier single-wavelength derivation [27] by treating wavelength explicitly. The corresponding squared interferometric visibility metric is

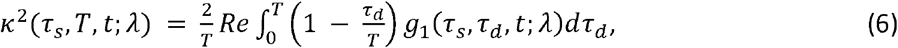

where T is the effective integration time, i.e., the upper limit of the retrospective numerical integration.

Equation (6) is the operative definition used in the present single-channel interferometric pipeline. The *κ*^2^ curve is reconstructed numerically from samples of *g*_1_, allowing *g*_1_-based and *κ*^2^-based analyses to be performed on the same interferometric record.

Importantly, Eq. (6) depends linearly on *Re*[*g*_1_], rather than on |*g*_1_|^2^. It therefore does not require a Siegert conversion. In the present implementation, the values of *T* define the upper limits of retrospective numerical integration. They do not represent physical camera exposures or averages of additional detector frames.

The same relation can be applied to the gate-resolved correlation 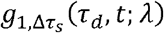 to obtain the corresponding gate-resolved visibility metric.

### 2.4. TOF- and wavelength-dependent decorrelation rate

Under a homogeneous-medium diffusing-wave-spectroscopy model with Brownian scatterer dynamics, the field autocorrelation at a fixed photon time-of-flight is approximated by a single exponential:

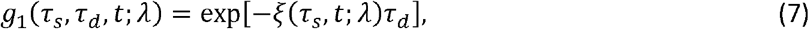

with TOF-dependent decorrelation rate

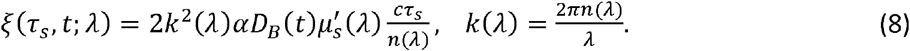

Here, *k(λ)* is the in-medium wavenumber, *αD*_*B*_ is the effective Brownian diffusion coefficient, conventionally used as a blood-flow index, 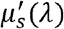 is the reduced scattering coefficient, and *n*(*λ*) is the refractive index.

Substitution of Eq. (7) into Eq. (6) gives

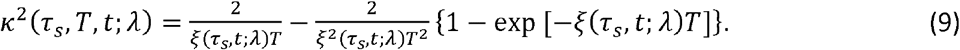

For fixed wavelength and homogeneous optical properties, *ξ* increases linearly with *τ*_*s*_. Later-arriving photons have, on average, traveled longer paths and undergone more dynamic scattering events. They therefore provide increased relative sensitivity to longer and typically deeper photon trajectories. Photon time-of-flight does not, however, map uniquely to anatomical depth.

For a finite TOF gate, an effective decorrelation rate 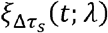 may be obtained by fitting the gate-resolved autocorrelation 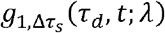 or 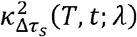. Because 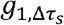, and 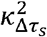 represent a mixture of photon path lengths, 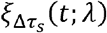 should be interpreted as an effective gate-level parameter.

Absolute decorrelation rates are not expected to match between 780 and 852 nm because 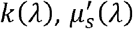 and the effective sampled dynamics can differ with wavelength [25]. Physiological changes are therefore normalized to the corresponding wavelength- and gate-specific baseline:

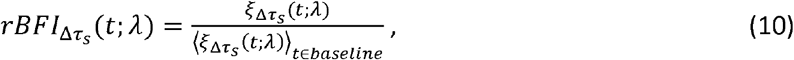

within a fixed wavelength and TOF gate, this relative decorrelation rate is used as an estimate of relative blood flow.

### 2.5 Spectral attenuation and two-chromophore inversion

The intensity branch is described using a differential, TOF-gate-resolved modified Beer-Lambert model. Whereas Sections 2.2-2.4 analyze temporal correlations of the recovered field, the spectroscopic branch uses the slow-time evolution of the TPSF itself.

For wavelength *λ* and TOF gate Δ*τ*_*s*_, the measured TPSF is integrated over the gate,

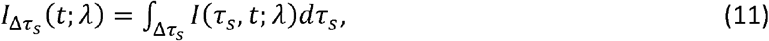

while the baseline TPSF is defined as

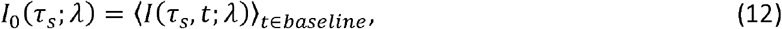

and the corresponding baseline gate intensity is

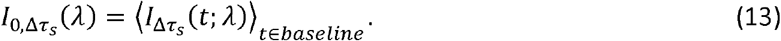

The gate-resolved change in optical density is then defined as

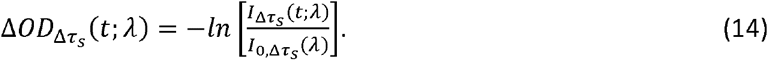

To relate gate-resolved *ΔOD* to absorption, we assume that differential attenuation is dominated by absorption and that scattering and the photon-path distribution remain approximately constant over the analyzed perturbation. For each wavelength and TOF gate, we first define the baseline TPSF-weighted mean photon time-of-flight as

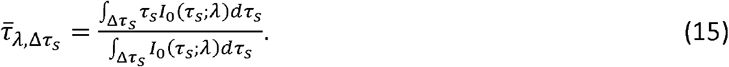

The quantity defined above represents the mean photon arrival time within the selected TOF gate, weighted by the baseline TPSF.

The corresponding effective mean photon path length is

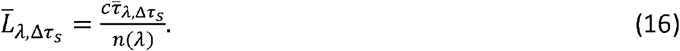

The gate-resolved change in absorption coefficient is then estimated using the differential modified Beer–Lambert law (MBLL),

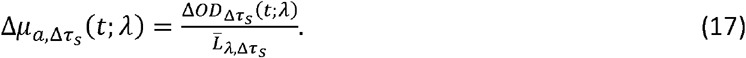

This expression provides a gate-wise differential MBLL estimate rather than an independently fitted absolute absorption coefficient. Each finite TOF gate contains a distribution of photon paths. Representing this distribution by a single mean path length does not isolate an anatomical layer or account for separate partial path lengths within different tissue compartments. Time-varying scattering, probe coupling, or motion may therefore bias the estimated 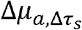.

The absorption changes at the two wavelengths are related to the chromophore concentration changes by

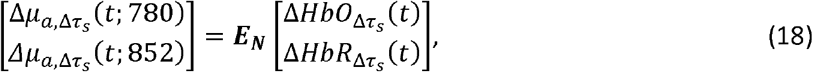

where ***E***_**N**_ is the natural-logarithm molar-extinction matrix of *HbO* and *HbR* at the two operating wavelengths. This inversion assumes that *HbO* and *HbR* are the dominant time-varying absorbers within this spectral interval, that their absorption contributions add linearly, that the wavelength channels are temporally aligned, and that the two-by-two extinction matrix is applicable to the sampled tissue.

The inversion yields the gate-resolved concentration changes *ΔHbO* and *ΔHbR*, with

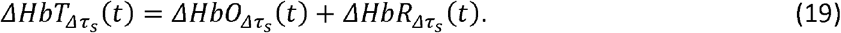

Because two wavelengths exactly determine two chromophore changes, wavelength-dependent path-length errors and measurement noise may appear as *HbO*/*HbR* cross-talk; absolute accuracy remains model dependent.

To estimate absolute tissue oxygen saturation, the differential chromophore changes must be referenced to baseline concentrations *BhO*_0_ and *HbR*_0_ for the selected TOF gate,

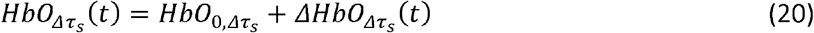

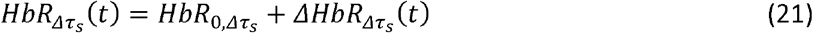

such that

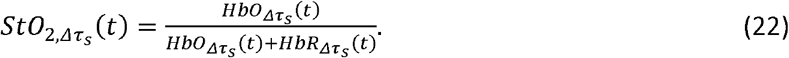

Absolute *StO*_2_ therefore cannot be determined from differential two-wavelength MBLL alone without an independently measured or assumed baseline chromophore state. The baseline values used in the present analysis are specified in Section 3.6.

### 2.6 Oxygen extraction and relative oxygen metabolic index

The oxygen-extraction fraction (OEF) is defined as the fraction of oxygen delivered in arterial blood that is extracted by tissue. Denoting arterial and venous oxygen contents by *C*_*aO*2_ and *c*_*v*_*O*_*2*_, respectively, and their corresponding hemoglobin oxygen saturations by *SaO* _*2*_ and *SvO*_*2*_,

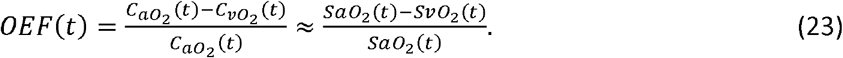

The saturation approximation neglects dissolved oxygen and assumes comparable hemoglobin concentration in the arterial and venous compartments.

Diffuse optical spectroscopy does not directly isolate *SvO*_*2*_. Instead, tissue oxygen saturation *StO*_2_ represents a path-weighted mixture of arterial, capillary, and venous compartments. If *StO*_2_ is treated as a venous-weighted surrogate, an effective gate-resolved oxygen-extraction estimate can be defined as

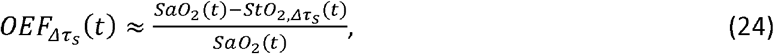

and its baseline-normalized form is

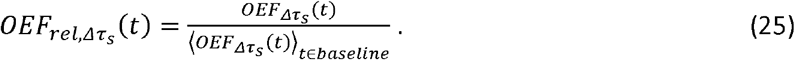

The resulting quantity should be interpreted as an optical surrogate of oxygen extraction rather than a direct measurement of venous OEF.

More generally, the Fick principle relates tissue oxygen consumption to blood flow, arterial oxygen content, and oxygen extraction,

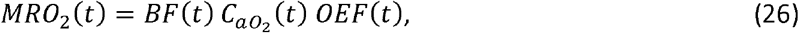

with the cerebral form commonly written as

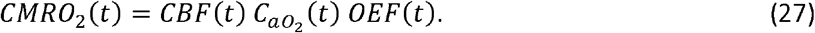

The present forearm measurements do not provide a direct measurement of *CMRO*_*2*_. Instead, under the assumptions that arterial oxygen content remains approximately constant and that *rBFI* is proportional to relative microvascular blood flow, we define a gate-resolved relative oxygen metabolic index as

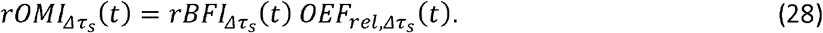

The *rOMI* quantity combines the relative microvascular-flow and oxygen-extraction responses and is therefore used here as a relative index of tissue oxygen consumption rather than as an absolute metabolic rate. Its interpretation requires sufficiently stable arterial oxygen content, proportionality between rBFI and relative microvascular flow, and sufficiently comparable tissue weighting of the flow and attenuation branches. Use of a common TOF gate and acquisition clock improves their correspondence but does not make their spatial sensitivities identical.

## 3. Materials and methods

### 3.1 Dual-wavelength swept-source interferometer

Two fiber-coupled distributed-feedback (DFB) lasers were used: a 780 nm source (EYP-DFB-0780-00020-1500-BFY12-0005, Eagleyard-Toptica) and an 852 nm source (EYP-DFB-0852-00015-1500-BFY12-0005, Eagleyard-Toptica). Their outputs were combined using a three-wavelength fiber wavelength-division multiplexer (WDM; RNN50HA, Thorlabs; nominal ports at 642, 785, and 852 nm; FC/APC connectors), with the 642 nm port unused, and routed through a shared Mach–Zehnder measurement interferometer (Fig. 2). Diffusely backscattered sample light was collected into a single-mode fiber and recombined with the reference field at a fiber coupler. The complementary coupler outputs were directed to a balanced detector. A portion of the reference arm light was also directed through a separate fixed-path reference interferometer, whose output provided the phase signal used for optical frequency linearization. The measurement and reference interferometer signals were digitized synchronously. Temporal wavelength multiplexing and wavelength assignment are described in Section 3.2. Table 1 summarizes the principal hardware and acquisition parameters, including the directly recorded and derived quantities used in the subsequent analysis. At the delivered powers of 11 mW at 780 nm and 9 mW at 852 nm, the calculated skin exposures remained approximately 2.5-fold and 4.3-fold below the respective maximum permissible exposure limits specified by ANSI Z136.1-2014.

**Table 1.**
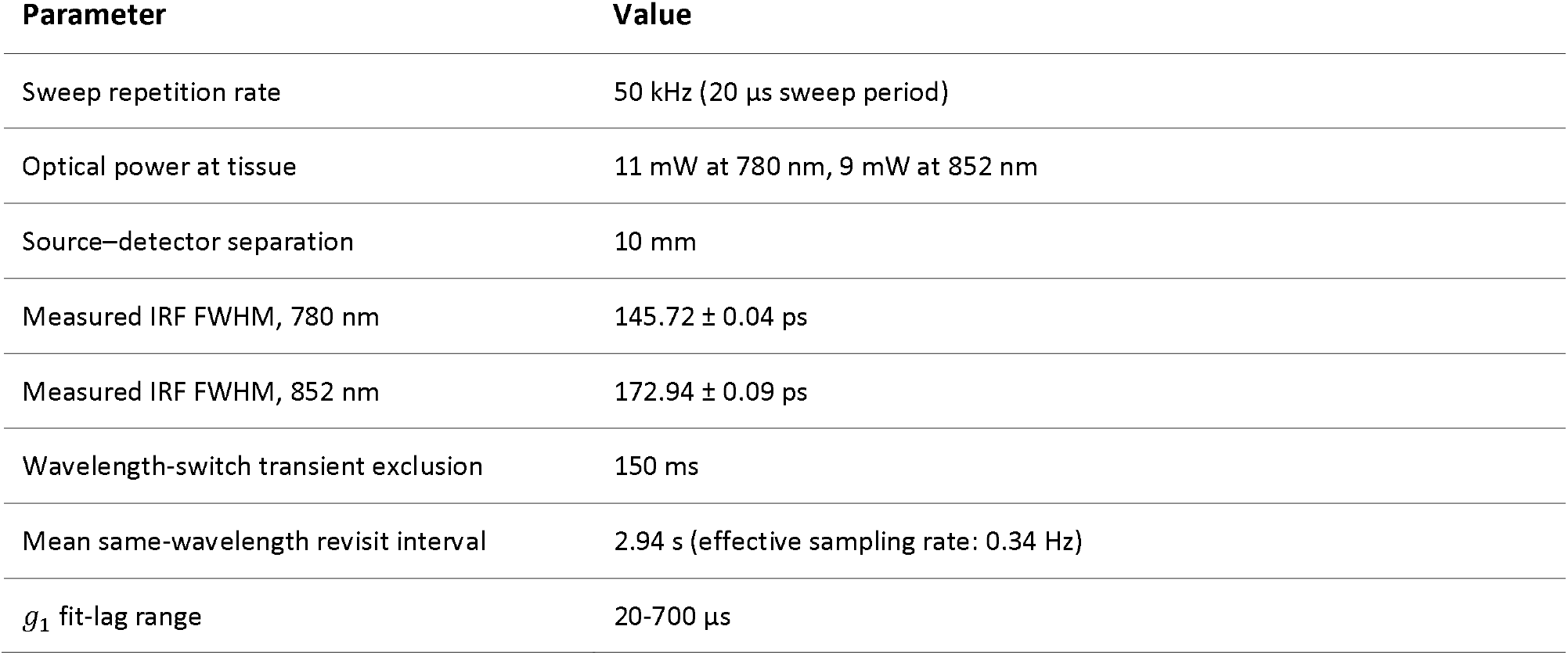
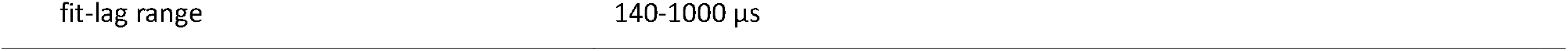
Acquisition and multiplexing parameters for the dual-wavelength TOF-iSCOS implementation.

**Table 2.** Participant characteristics. Continuous variables are reported as mean ± SD (range). Categorical variables are reported as counts. One participant contributed bilateral recordings, which were averaged before participant-level inference. BMI, body mass index.

| Characteristic | Value |
| --- | --- |
| Independent participants | 11 |
| Sex | 9 male, 2 female |
| Age | 36.6 ± 6.8 years; median 40 years; range 24-44 years |
| BMI | 25.3 ± 3.3 kg m<sup>-2</sup>; median 24.2 kg m<sup>-2</sup>; range 21.0-32.0 kg m<sup>-2</sup> |
| Fitzpatrick skin type | median 2; range 2-5; type II n=8, type III n=1, type IV n=1, type V n=1 |

**Figure 2.**
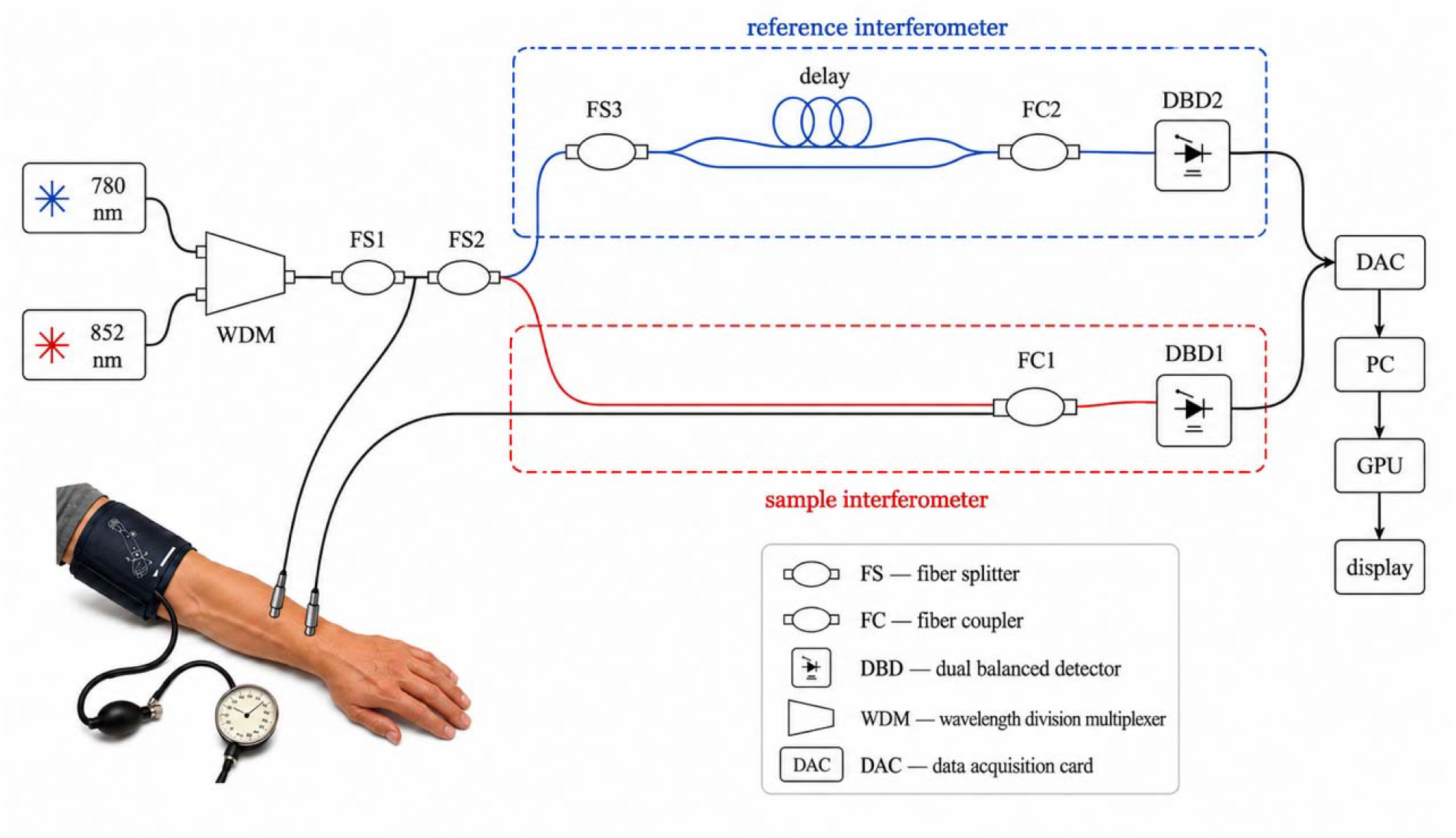
Dual-wavelength optical layout and forearm cuff geometry. (A) The 780 and 852 nm swept sources are combined with a three-wavelength WDM (RNN50HA, Thorlabs; 642-nm port unused) and routed through the shared measurement interferometer. Measurement and k-clock/reference interferometers are detected by balanced detectors and digitized synchronously by a three-channel data-acquisition card (DAC). (B) Forearm cuff geometry with the optical probe placed on the ventral forearm at 10-mm source-detector separation. Abbreviations: WDM, wavelength-division multiplexer; DAC, data-acquisition card.

### 3.2 Temporal wavelength multiplexing

Measurements at 780 and 852 nm were performed within a single alternating-wavelength session composed of sequential wavelength-specific acquisition chunks (Fig. 3). Each laser was driven by a dedicated generator (DG1032Z, Rigol), and only one source was swept at a time. During acquisition at one wavelength, the waveform for the other wavelength was configured in preparation for the subsequent chunk. Wavelength identity was therefore assigned prospectively from the acquisition sequence and stored together with the corresponding acquisition timestamps; no post hoc demultiplexing of a simultaneously recorded mixed-wavelength signal was required.

**Figure 3.**
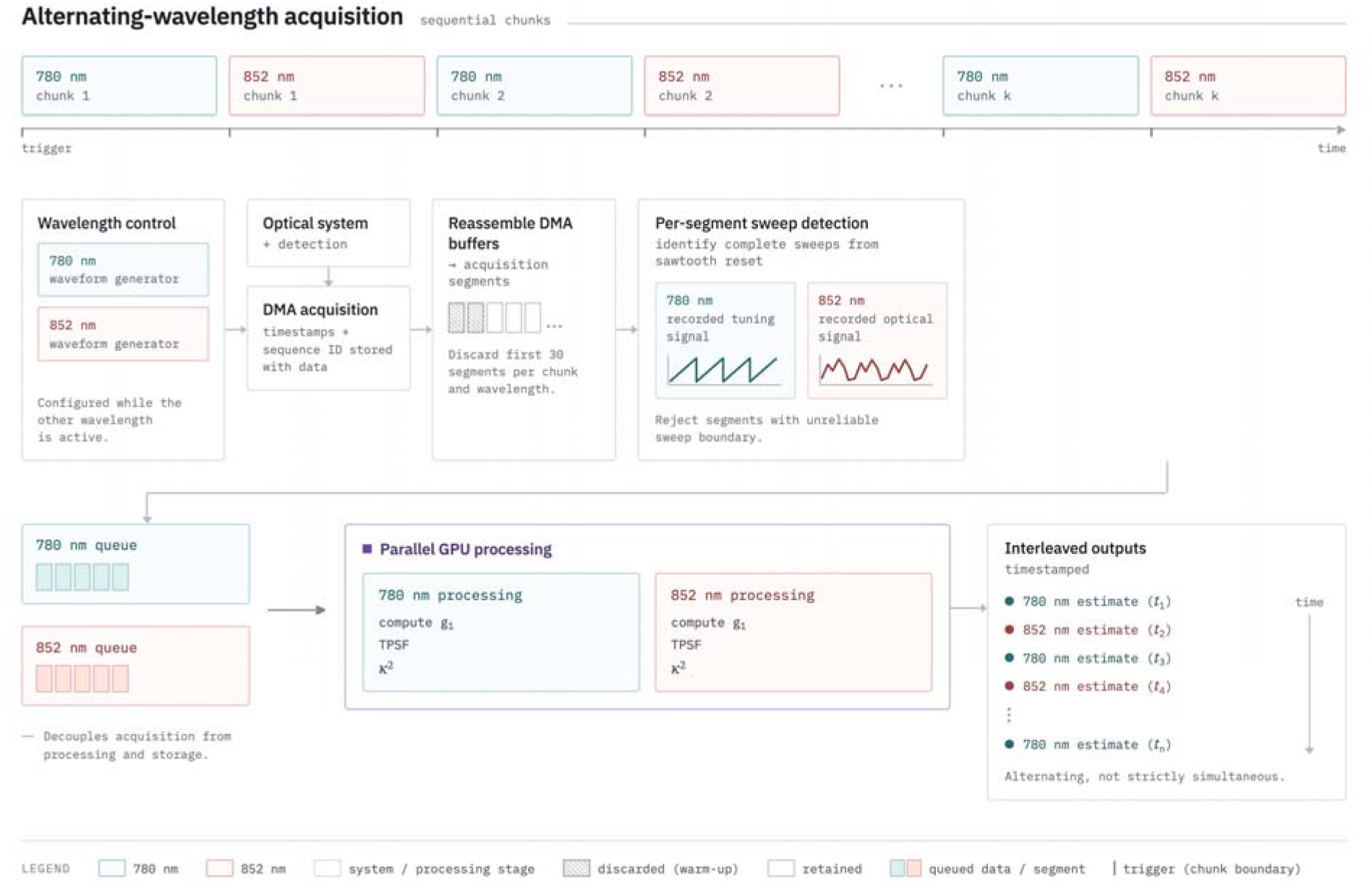
Alternating-wavelength acquisition and parallel processing workflow. The 780 and 852 nm sources are acquired in sequential chunks, with trigger events marking the chunk boundaries; while one wavelength is active, the waveform generator for the other is configured. Digitizer data are reassembled from direct-memory-access (DMA) buffers into processing segments, and complete sweeps are identified using wavelength-specific boundary detection: the recorded tuning signal at 780 nm and the optical signal at 852 nm. Segments without reliable sweep boundaries are discarded. Accepted segments are placed in asynchronous wavelength-specific queues and processed in parallel by persistent GPU contexts to compute autocorrelation and the TPSF (the visibility factor is computed in post processing). The timestamped wavelength-specific estimates are subsequently written to an interleaved output stream. Green and red denote the 780 and 852 nm channels, respectively. The remaining colors and fill patterns distinguish acquisition, processing, retained output and discarded data.

Signals were acquired using a Spectrum M2p.5962-x4 16-bit digitizer operated at 100 MS/s per channel. The sweep repetition rate was 50 kHz, corresponding to a sweep period of 20 µs and 2000 digitizer samples per sweep. The continuous data stream was reassembled into logical processing segments containing approximately 250 sweeps. After trigger and buffer alignment, each segment contained 500,224 samples per channel and represented approximately 5.0 ms of acquired data.

To suppress transients associated with wavelength switching and laser-tuning stabilization, the first 30 segments following each wavelength transition were excluded from processing, corresponding to approximately 150 ms. Complete sweeps were subsequently identified within each retained segment. Sweep-boundary detection was wavelength specific: the recorded tuning signal was used at 780 nm, whereas the optical signal was used at 852 nm. Segments for which reliable sweep boundaries could not be identified were rejected.

Accepted segments were dispatched to two persistent GPU processing contexts, one for each wavelength. Processing was performed asynchronously with acquisition, and completed results were transferred to a separate storage queue. Acquisition of the subsequent wavelength could therefore begin without waiting for processing or disk writing to finish.

Within each accepted segment, the first-order field autocorrelation was calculated from mean-subtracted complex Fourier fields of consecutive sweeps. For correlation lag m, N-m field pairs contributed to the estimate, where N was the number of complete sweeps in the segment. Segment-level correlation matrices were averaged over the retained portion of each wavelength chunk and normalized by their lag-zero values. The maximum evaluated correlation lag was determined by the configured lag range and the number of complete sweeps within an individual segment rather than by the total chunk duration.

In a 307-chunk timing validation, the mean same-wavelength revisit interval was approximately 2.94 s, corresponding to an effective wavelength-specific sampling rate of approximately 0.34 Hz. The wavelength-to-wavelength transition intervals were asymmetric because waveform preparation and acquisition overhead differed between the two directions, but processing remained faster than the revisit interval and did not accumulate a backlog. The 780 and 852 nm estimates were therefore temporally interleaved and registered to their acquisition timestamps, but were not strictly simultaneous.

Three 16-bit channels sampled at 100 MS/s generated a raw data rate of approximately 4.8 Gbit/s, corresponding to approximately 0.60 GB per nominal 1s acquisition chunk. The processed representation, containing reconstructed TOF profiles, correlation matrices, coordinate axes, processing metadata, and acquisition timestamps, required approximately 0.58 MB per chunk, corresponding to a reduction in stored data volume of approximately three orders of magnitude. For the 307-chunk validation sequence, this corresponds to approximately 185 GB of raw data compared with approximately 179 MB of processed data.

### 3.3 Wavelength-specific TOF reconstruction and gating

The 780 and 852 nm data streams were reconstructed independently using wavelength-specific spectral linearization and TOF calibration (Fig. 4). Within each accepted sweep, the usable spectral interval was selected and resampled onto a uniform optical-frequency, equivalently uniform wavenumber, grid using the phase of the corresponding reference-interferometer signal. The resampled interferograms were detrended, mean-subtracted, apodized, zero-padded, and Fourier transformed to recover the complex delay-resolved optical field. Only the positive TOF branch was retained.

**Figure 4.**
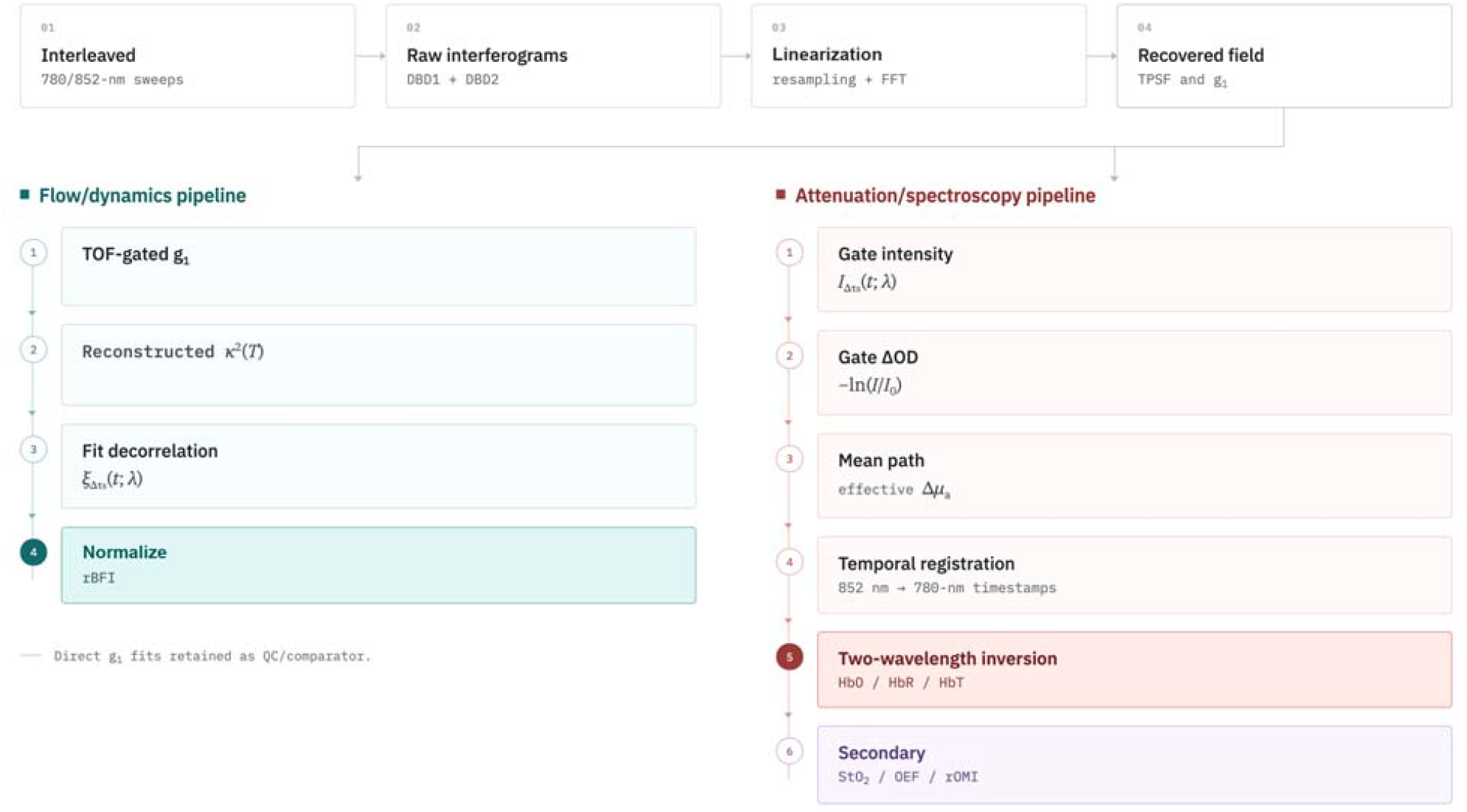
Dual-wavelength TOF-iSCOS processing workflow. Interleaved 780 and 852 nm sweeps are detected as balanced raw interferograms, linearized in optical frequency, resampled, and Fourier transformed to recover the complex field, TPSF, and field autocorrelation *g*_l_. The recovered data are then analyzed through two parallel pipelines. The flow/dynamics pipeline calculates TOF-gated *g*_1_, reconstructs the finite-integration visibility *κ*^2^*(r)*, fits the decorrelation rate 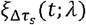, and normalizes it to obtain relative blood-flow index (rBFI); direct *g*_l_ fits are retained as a quality-control comparator. The attenuation/spectroscopy pipeline converts gate-integrated intensity into changes in optical density and effective absorption, temporally registers the wavelength channels, and performs two-wavelength inversion to recover *ΔHb0, ΔHbR*, and *ΔHbT*. Secondary oxygenation metrics include *StO*_2_, oxygen-extraction fraction (OEF), and the relative oxygen metabolic index (rOMI).

A reference-interferometer path length of 0.5 m was used for TOF calibration. The reconstructed TOF-bin spacing was 12.62 ps at 780 nm and 14.97 ps at 852 nm. The measured wavelength-specific instrument-response-function (IRF) widths were 145.72 ± 0.04 ps at 780 nm and 172.94 ± 0.09 ps at 852 nm. The TOF-bin spacing therefore represents numerical sampling of the reconstructed axis and should not be interpreted as the physical temporal resolution, which was determined from the measured IRF width.

The raw Fourier-delay coordinate was calibrated independently for each wavelength. The wavelength-specific instrument response was measured under direct source-to-collection coupling with appropriate attenuation, and the position of its peak defined the zero-delay reference. No IRF deconvolution was applied. The reconstructed TOF profiles and derived path-length quantities therefore remained instrument-convolved.

For subsequent analysis, the wavelength-specific TOF axes were registered to a common calibrated coordinate system. Path-length calculations assumed a tissue refractive index of 1.4. The cohort analysis used three 50 ps TOF windows fixed before group-level inference: Early, 75–125 ps; Middle, 325–375 ps; and Late, 525–575 ps. Early sampled photons preceding the TPSF maximum, Middle sampled the early falling edge, and Late sampled the later TPSF tail. These windows were selected from development-data gate and fit-range analyses and then kept fixed for all 12 cohort recordings. Gate-dependent effects were interpreted as changes in overlapping photon-path populations rather than isolation of discrete anatomical layers.

The processed output retained the wavelength-specific TPSF and IRF, TOF and correlation-lag axes, the TOF-resolved field-correlation matrices, and the reconstructed Fourier-magnitude profiles required for subsequent flow and spectroscopic analyses.

### 3.4 TOF-resolved flow estimation

The primary dynamic observable was the wavelength- and TOF-resolved first-order field autocorrelation *g*_1_, calculated directly from the reconstructed complex optical fields. For each TOF gate, the retained segment-level correlations were combined to obtain a gate-resolved *g*_1_ curve, from which the TOF-dependent decorrelation rate *ξ* was estimated.

Two related estimator routes were evaluated. In the g_1_-based estimator, the decorrelation rate 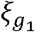 was obtained by fitting the gate-resolved *g*_1_ decay with the single-exponential model of Eq. (7). In the *κ*^2^-based estimator, the finite-integration visibility metric *κ*^2^ was reconstructed numerically from the measured *g*_1_ samples using Eq. (6), and the decorrelation rate 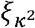 was obtained by fitting the corresponding analytical model of Eq. (9). The zero-lag sample was excluded from both fitting routes to reduce sensitivity to the local zero-lag artifact.

The *g*_1_-based and *κ*^2^-based routes originate from the same interferometrically measured field correlation and are therefore not statistically independent estimators. They differ, however, in their numerical transformation and fitting domains. The *κ*^2^-based estimator was used as the primary operational flow estimate because it provided a higher fraction of accepted estimates in the lower-photon late gates, whereas the *g*_1_-based estimator was retained as a confirmatory analysis. This difference in accepted fraction should be interpreted as an operational robustness difference within the present processing pipeline rather than as an intrinsic estimator-efficiency advantage.

Fits were accepted only when the estimated decorrelation rate remained within the predefined physiological fitting bounds and the coefficient of determination satisfied *R*^2^ ≥ 0.80. Relative blood-flow indices were obtained by normalizing accepted *ξ* estimates to the corresponding wavelength- and gate-specific baseline, as defined in Eq. (10). For the physiological experiments, the baseline was calculated over the predefined pre-perturbation interval specified below.

As a baseline optical-property consistency check, the baseline TPSFs from the 11-participant cohort were fitted to a semi-infinite time-domain diffusion model to estimate effective *μ*_*a*_ and 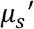 t 780 and 852 nm. These estimates document the effective optical-property scale for the 10 mm SDS forearm geometry but were not used for cohort-level inference or for converting the reported rBFI and differential MBLL outcomes into absolute quantities (Supplementary Fig. S1).

Agreement between the *g*_1_ and *κ*^2^ processing routes was evaluated in terms of covariation, preservation of physiological responses, and the relationship between their estimated decorrelation rates. Because both estimators were derived from the same measured field autocorrelation, this comparison was treated as an internal processing cross-check rather than validation against an independent blood-flow measurement.

### 3.5 Relative blood-flow index and spectroscopic quantities

For each wavelength and TOF gate, accepted decorrelation-rate estimates were converted to relative blood-flow index (rBFI) by normalization to the corresponding mean value over the 0.5–2.0 min baseline interval,

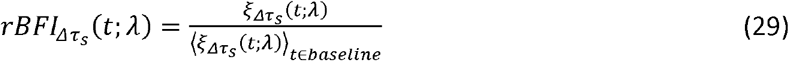

Absolute *αD*_*B*_ was not used for cohort-level inference because its estimation requires wavelength-specific optical properties and a more complete instrument and tissue model.

For the spectroscopic branch, the TPSF intensity was integrated within each fixed TOF gate and referenced to the corresponding 0.5-2.0 min baseline. The mean TPSF over this baseline interval was used as the baseline TPSF *I*_0_(*τ*;*λ*), consistent with the definition in Section 2.5. Gate-resolved optical-density changes were calculated as −ln(*I*/*I*_0_) and converted to effective absorption changes using the baseline TPSF-weighted mean photon path length. The resulting quantities are therefore effective gate-resolved Δ*µ*_*a*_ values in cm^-1^, rather than independently fitted absolute absorption coefficients.

Because the 780 and 852 nm measurements were acquired sequentially rather than simultaneously, the wavelength-specific *Δµ*_*a*_ traces were temporally registered using their acquisition timestamps before spectroscopic inversion. The 852 nm traces were linearly interpolated onto the 780 nm time grid, and the two-wavelength inversion was performed only over time points for which both wavelength estimates were available. No extrapolation beyond the common temporal support was used.

The aligned absorption-change vectors were converted to oxy- and deoxyhemoglobin concentration changes using the natural-logarithm molar-extinction matrix [Eq. (18)]. The extinction coefficients were obtained from the Prahl spectra compiled by the Oregon Medical Laser Center (OMLC) and converted from base-10 to natural-logarithm form by multiplication by ln(10) [30]. The resulting matrix was:

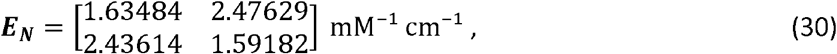

with rows corresponding to 780 and 852 nm and columns to HbO and HbR, respectively. Total hemoglobin change was calculated using Eq. (19). For estimation of absolute tissue oxygen saturation, baseline total hemoglobin concentration and tissue saturation were assumed to be HbT_0_=0.119 mM and StO_2_,_0_=0.73, respectively, based on resting forearm-muscle values reported by Re et al. [31]. These values correspond to

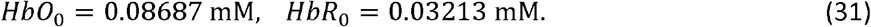

Absolute chromophore concentrations were reconstructed using Eqs. (20)-(21) and tissue oxygen saturation was calculated with Eq. (22). Arterial oxygen saturation was assumed to remain constant during the experiment. The effective gate-resolved oxygen-extraction fraction was therefore estimated according to Section 2.6 and normalized to its baseline value,

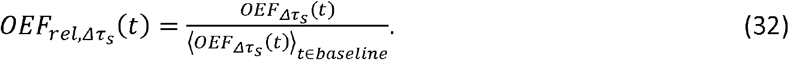

Finally, relative oxygen metabolism was represented by the relative oxygen metabolic index,

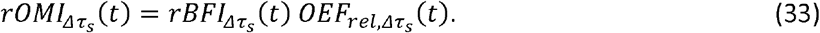

The OEF and rOMI quantities were treated as secondary, model-derived within-participant outcomes. They should be interpreted as relative optical indices of oxygen extraction and oxygen consumption rather than as direct measurements of venous oxygen saturation or absolute tissue metabolic rate. No physical-range clipping was applied.

For calculation of response metrics, rBFI trajectories were smoothed using a three-frame moving median followed by a three-frame moving mean, whereas attenuation-derived trajectories were smoothed using a three-frame moving mean. Additional smoothing used for visualization was applied only after participant-level quantities had been calculated. Scalar response metrics and inferential statistical tests were based on the corresponding unsmoothed trajectories.

### 3.6 Forearm cuff protocol and participants

Eleven healthy adults contributed 12 uninterrupted forearm recordings. Participant P01 was measured on both arms; these recordings were retained separately for session-level quality control and averaged before participant-level statistical inference.

The optical probe was positioned on the ventral forearm, with a pneumatic cuff placed on the ipsilateral upper arm above the elbow. After a 2.5 min baseline period, the cuff was inflated to 180 mmHg and maintained for 2.0 min, followed by cuff release and post-occlusion monitoring to a total recording duration of approximately 7.5 min.

All recordings were processed using the same predefined TOF gates, fitting procedures, quality-control thresholds, baseline interval, and response definitions. Participants provided written informed consent. All procedures were approved by the Commission of Bioethics at the Military Institute of Medicine, Poland, permission no. 90/WIM/2018, and were conducted in accordance with the Declaration of Helsinki.

A separate hypothesis-directed control measurement was subsequently acquired in participant P01 to examine the effect of a longer occlusion period. This recording used the same instrumentation, probe geometry, processing pipeline, and TOF gates, but extended cuff occlusion to 3.5 min. The control recording was analyzed separately, was not used to modify the processing pipeline or analysis definitions, and was excluded from all cohort-level summaries and inferential statistics.

### 3.7 Statistical analysis

All response quantities were first calculated independently for each recording. The two arm recordings from participant P01 were subsequently averaged at participant level to avoid pseudoreplication. Group trajectories and phase summaries are reported as median and interquartile range (IQR), with individual participant responses retained where appropriate. Inferential analyses were performed using participant-level values.

Baseline was defined as 0.5-2.0 min. Cuff onset and release were determined from the temporal derivative of the median 780 nm rBFI response across TOF gates, using predefined search intervals surrounding the expected inflation and release times. The stable-occlusion window excluded the initial transient following cuff inflation and the final minute preceding release, whereas the reactive-hyperemia window extended for 45 s following cuff release.

For estimator *e* and TOF gate *Δτ*_*s*_, cuff-induced flow suppression was defined as

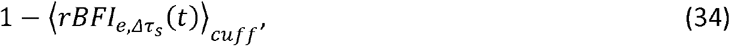

and reactive-hyperemia amplitude as

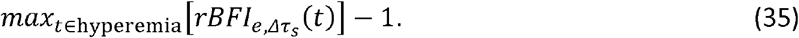

The attenuation response was defined as the mean 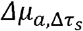 over the stable-occlusion interval. Flow-response metrics were retained only when at least 70% of fitted estimates within the corresponding response interval satisfied the predefined *R*^2^ ≥ 0.80 quality criterion.

Primary gate contrasts were prespecified as Late-minus-Early for cuff-induced rBFI suppression and reactive hyperemia, and Early-minus-Late for the attenuation response. Gate ratios were not used as primary statistical outcomes.

Hemoglobin responses were additionally characterized using predefined temporal phases relative to cuff inflation and release. Early post-inflation responses were quantified over the first 20 s following cuff onset, subsequent occlusion trends were characterized by linear regression during the sustained-cuff period, and end-cuff levels were calculated over the final 20 s before release. Early reperfusion slopes and post-release extrema were quantified over predefined windows following cuff release. The same temporal definitions were applied to all participants and TOF gates without adjustment according to the observed direction of the HbO or HbR response.

Participant-level paired contrasts were summarized using 10,000 bootstrap resamples. Prespecified directional gate hypotheses were tested using exact one-sided Wilcoxon signed-rank tests. Exploratory phase-resolved hemoglobin Early-to-Late contrasts, for which no directional sign was prespecified, were evaluated using exact two-sided Wilcoxon signed-rank tests. Because the tested contrasts represented a small number of predefined directional hypotheses for each observable, reported P values were not adjusted for multiple comparisons. Directional consistency across participants was additionally summarized using exact one-sided binomial sign tests against a probability of 0.5.

Agreement between the *g*_1_-based and *κ*^*2*^-based decorrelation-rate estimates, 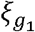 and 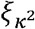, was evaluated using Pearson correlation on the linear rate scale and complementary log–log regression. Confidence intervals were obtained by participant-level cluster bootstrap resampling. Accepted-frame fractions were summarized by wavelength and TOF gate as operational measures of estimator robustness, rather than as a controlled comparison of estimator efficiency.

One representative recording is shown for visualization of the processing and physiological responses. The recording was selected before figure preparation from sessions passing completeness quality control as the session with stable *κ*^*2*^ fits at both wavelengths and response amplitudes closest to the cohort medians across the main rBFI and *Δµ*_*a*_ metrics. This illustrative choice did not affect any cohort-level analysis or statistical inference.

## 4. Results

### 4.1 One multiplexed record resolves flow and attenuation at two wavelengths

All 12 recordings were reconstructed without restarting the interferometer. Figure 5 first provides the TPSF context for the fixed Early, Middle, and Late TOF gates (Fig. 5A). In illustrative session P08-L, rBFI at both wavelengths was stable during baseline, fell rapidly after cuff inflation and showed reactive hyperemia after release (Fig. 5B,C). The Late-gate overshoot exceeded the Early and Middle responses. In the same record, effective *Δµ*_*a*_ increased during occlusion at both wavelengths, with the largest response in Early and the smallest in Late (Fig. 5D,E). Thus, one multiplexed acquisition recovered the imposed vascular transition through two observables with opposite TOF weighting.

**Figure 5.**
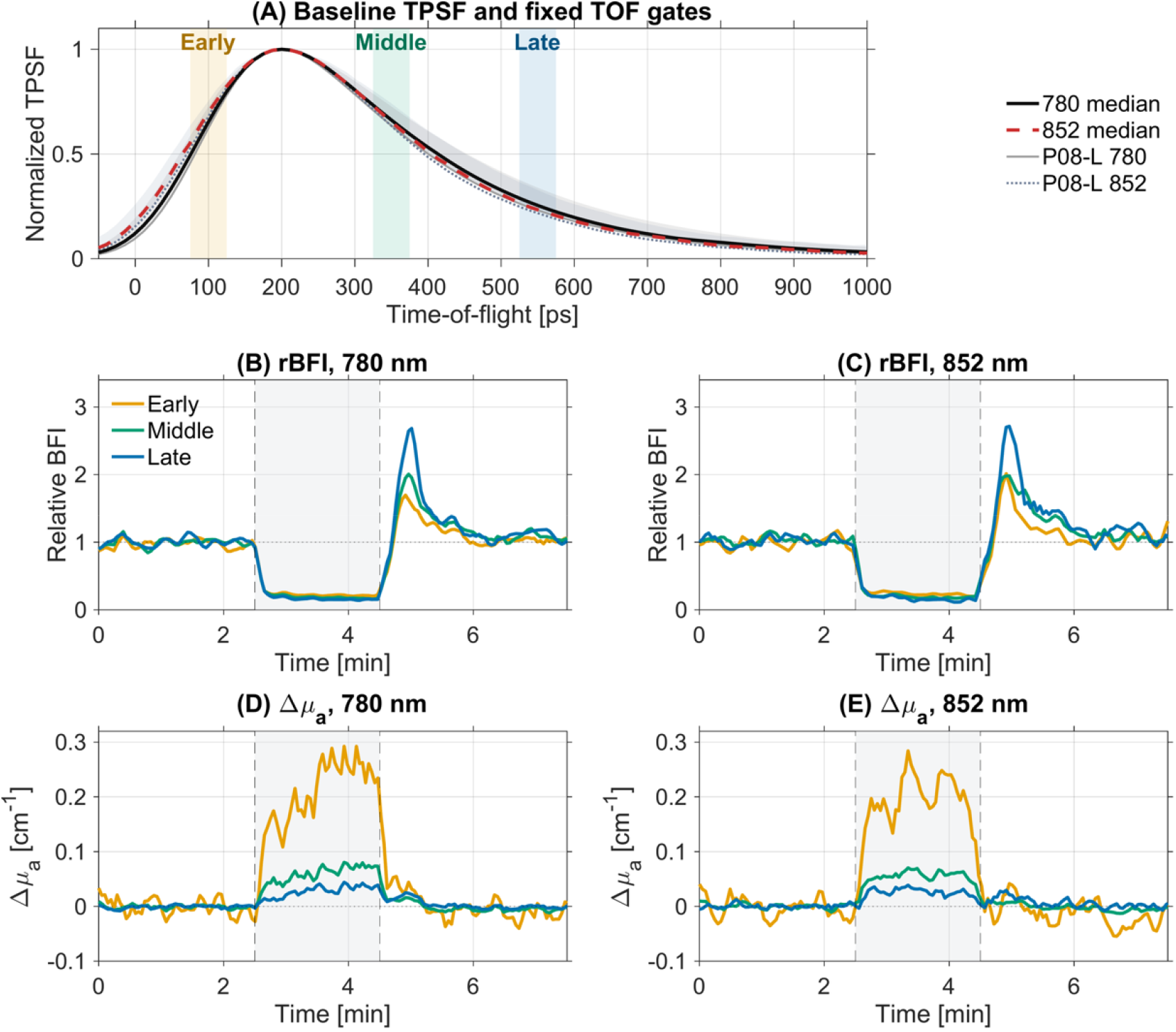
Illustrative co-registered multi-contrast record with TPSF gate orientation for selected participant (P08-L). (A) Baseline normalized TPSFs show the cohort median with interquartile range across 11 independent participants, with P08-L shown as thin traces; shaded bands mark the Early (75-125 ps), Middle (325-375 ps) and Late (525-575 ps) TOF gates. (B,C) *κ*^2^-based rBFI at 780 and 852 nm. (D,E) Gate-resolved effective *Δµ*_*a*_ at 780 and 852 nm. The gray band denotes cuff occlusion; dashed vertical lines mark cuff inflation and release. Gate colors are shared across the time-course panels.

### 4.2 *K*^2^-based fitting retains higher late-gate fit yield than *g*_*1*_-based fitting

Rejected *g*_1_-based fits were concentrated in the photon-limited 852 nm Late gate (Fig. 6A,B). Across participants, the median accepted-frame fraction in this gate was 0.831 for the *g*_*1*_-based estimator (minimum 0.000) and 0.994 for the *K*^*2*^-based estimator (minimum 0.928). The *κ*^*2*^-based estimator retained at least 92.8% of frames in every participant, wavelength and gate combination, whereas P11-L had no accepted *g*_*1*_-based frames in the 852 nm Late gate. At the stage-averaged level, 214 of 216 wavelength-, gate- and stage-resolved pairs met both estimators’ acceptance criteria.

**Figure 6.**
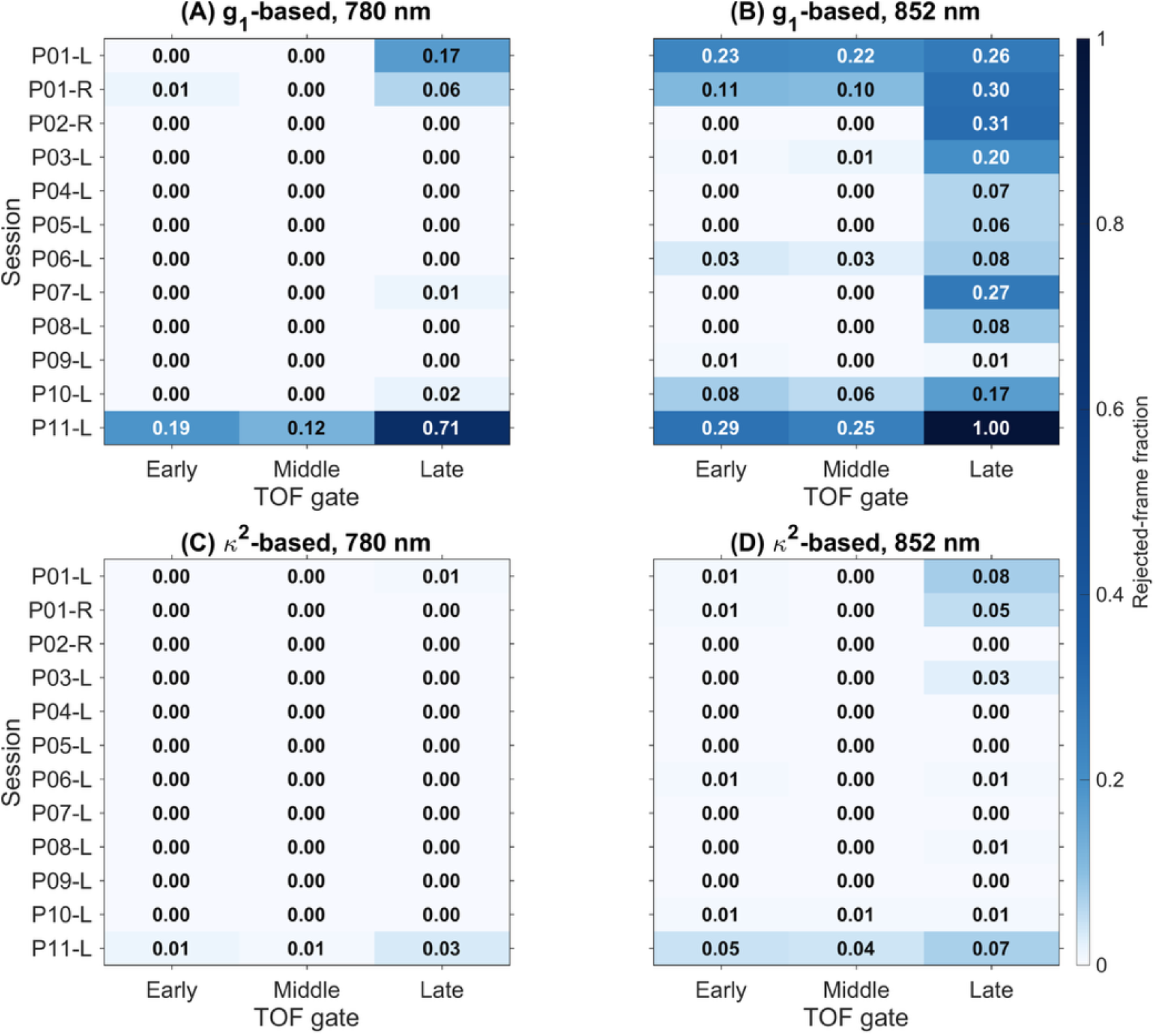
Gate- and wavelength-resolved estimator stability across 12 sessions. Square heat maps show the rejected frame fraction, 1 minus the accepted fraction, for direct *g*_*1*_ fits and reconstructed *κ*^2^ fits at 780 and 852 nm. Columns indicate Early, Middle, and Late TOF gates, and rows indicate pseudonymized recording session codes. Values above the common 0-0.35 color scale are printed in white. The *κ*^2^ estimator reduces late-gate rejections, especially at 852 nm.

This comparison motivates use of the *κ*^*2*^-based estimator as the primary operational flow estimator for the cohort. It does not establish intrinsic estimator superiority: *κ*^*2*^ is integrated from positive-lag *g*_*1*_, its fit domain differs, and integral smoothing makes the *R*^*2*^ threshold easier to satisfy. Detailed rate agreement and bias analyses are reported in Supplementary Fig. S3.

### 4.3 Flow and attenuation show distinct TOF-gate weighting across participants

Participant-level time courses reproduced the illustrative cuff response at both wavelengths (Fig. 7). rBFI decreased promptly after cuff inflation and showed reactive hyperemia after release, with the largest post-release response occurring in the Late gate. Effective Δ*µ* increased during occlusion and returned toward baseline after release but followed the opposite TOF ordering: the response was largest in Early and smallest in Late. The same qualitative patterns were observed at 780 and 852 nm.

**Figure 7.**
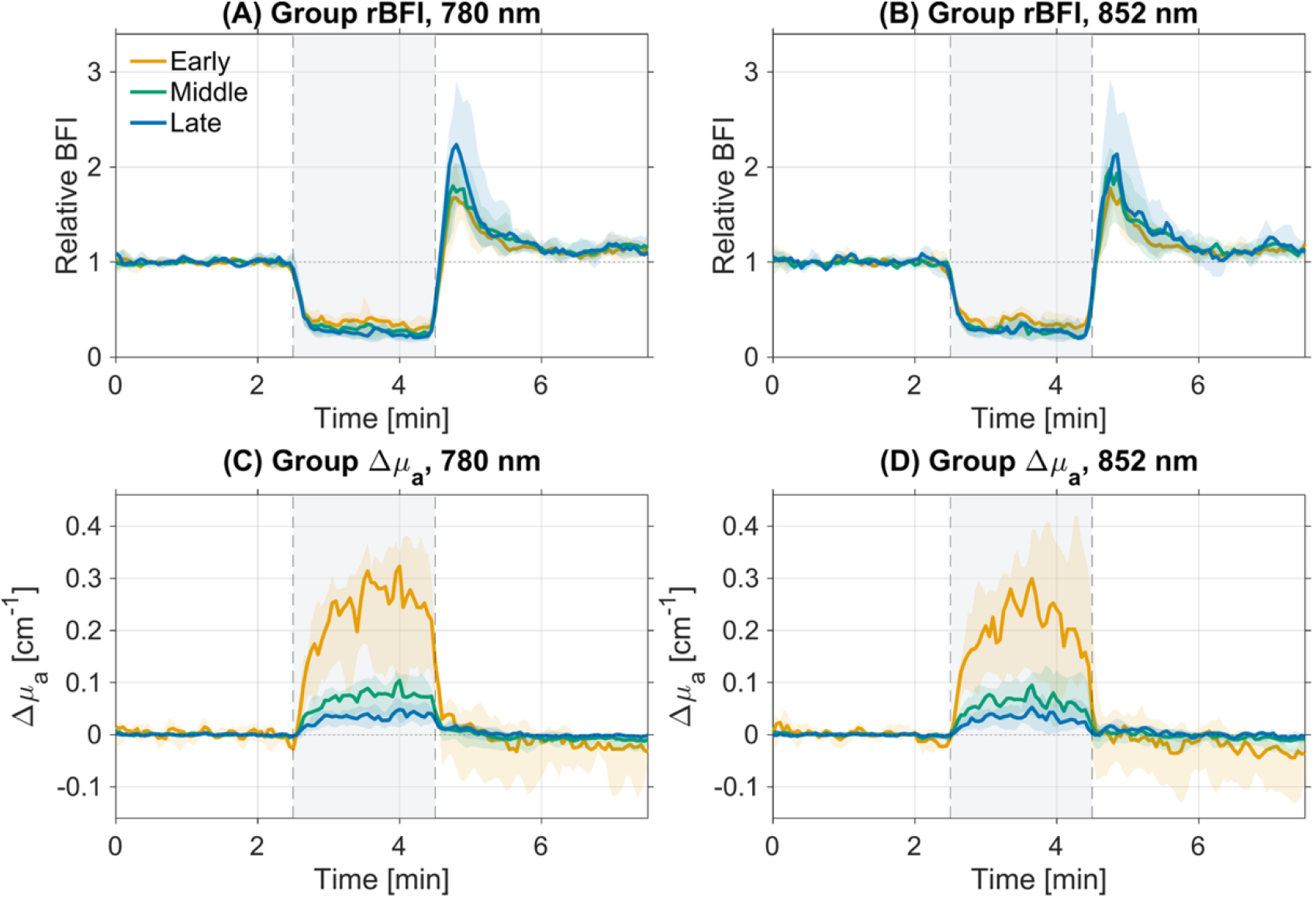
Group TOF-resolved flow and attenuation time courses during forearm cuff occlusion (N = 11 independent participants). (A,B) *κ*^2^-based relative blood-flow index (rBFI) at 780 and 852 nm, respectively. (C,D) Effective absorption-coefficient changes, Δ*µ*_*a*_, at 780 and 852 nm, respectively. Solid colored curves show participant medians, and color-matched shaded bands show the interquartile range (IQR). Orange, green, and blue denote the Early, Middle, and Late TOF gates. The gray background marks cuff occlusion, and vertical dashed lines indicate nominal cuff inflation and release.

These group trajectories demonstrate that the TOF dependence is not restricted to a representative recording. Later-arriving photons preferentially emphasized the post-release flow response, whereas early-arriving photons carried the strongest attenuation response during occlusion. Flow-sensitive field dynamics and wavelength-resolved attenuation therefore provide complementary TOF-dependent contrasts within the same acquisition.

Two-wavelength inversion of the gate-resolved attenuation traces revealed distinct hemoglobin dynamics across photon time-of-flight (Fig. 8). The Early gate showed the largest increases in ΔHbO, ΔHbR, and ΔHbT during cuff occlusion, whereas the Middle and Late responses were smaller. OEF increased during occlusion, consistent with increased fractional oxygen extraction under restricted flow. The *κ*^*2*^-based rOMI was strongly suppressed during occlusion and exhibited a post-release overshoot across all three TOF gates.

**Figure 8.**
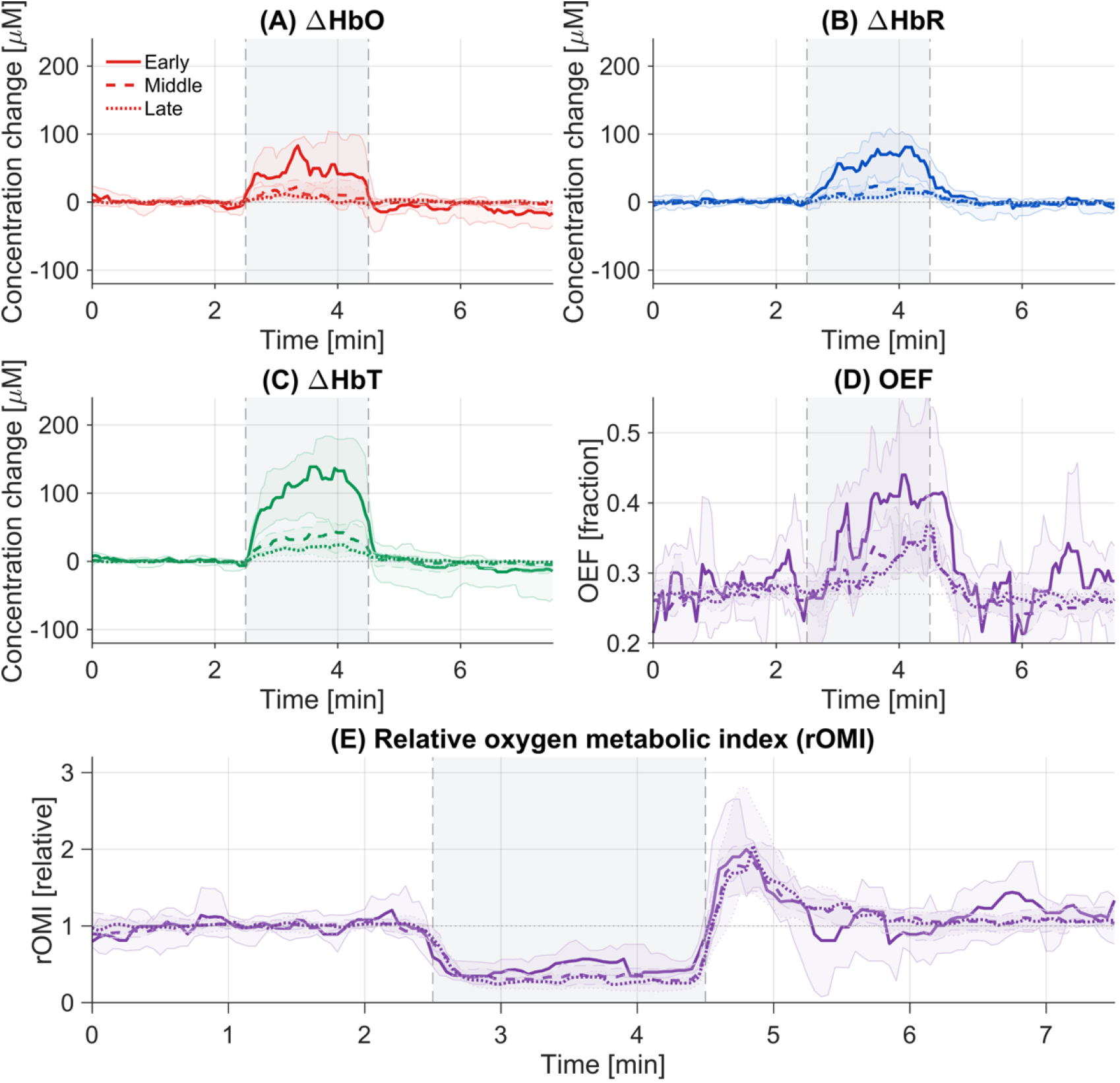
Gate-resolved hemoglobin, oxygen-extraction and relative metabolic trajectories (N = 11 independent participants). (A–C) Changes in oxyhemoglobin, deoxyhemoglobin, and total hemoglobin concentration derived from the two-wavelength attenuation branch and expressed in µM. (D) Model-derived oxygen-extraction fraction (OEF). (E) Relative oxygen metabolic index (rOMI), calculated as the product of *κ*^2^-based relative blood flow and relative OEF. Solid, dashed and dotted lines represent the Early, Middle and Late TOF gates, respectively. Curves and color-matched shades show participant median and IQR. The gray region denotes cuff occlusion from 2.5 to 4.5 min; dashed vertical lines mark cuff inflation and release. Participant trajectories were smoothed using a seven-frame moving median before group summarization. Bilateral recordings from P01 were averaged before cohort aggregation. OEF and rOMI are model-dependent relative observables based on literature-assigned baseline hemoglobin values.

### 4.4 Response amplitude and physiological phase diverge across TOF gates

The complete trajectories also indicated that the hemoglobin response was not temporally uniform: the initial vascular-volume change and the subsequent evolution during sustained occlusion differed across TOF gates. To quantify this behavior, the response was separated into an initial 20 s interval after cuff inflation and a sustained-cuff interval extending to 10 s before release (Fig. 9).

**Figure 9.**
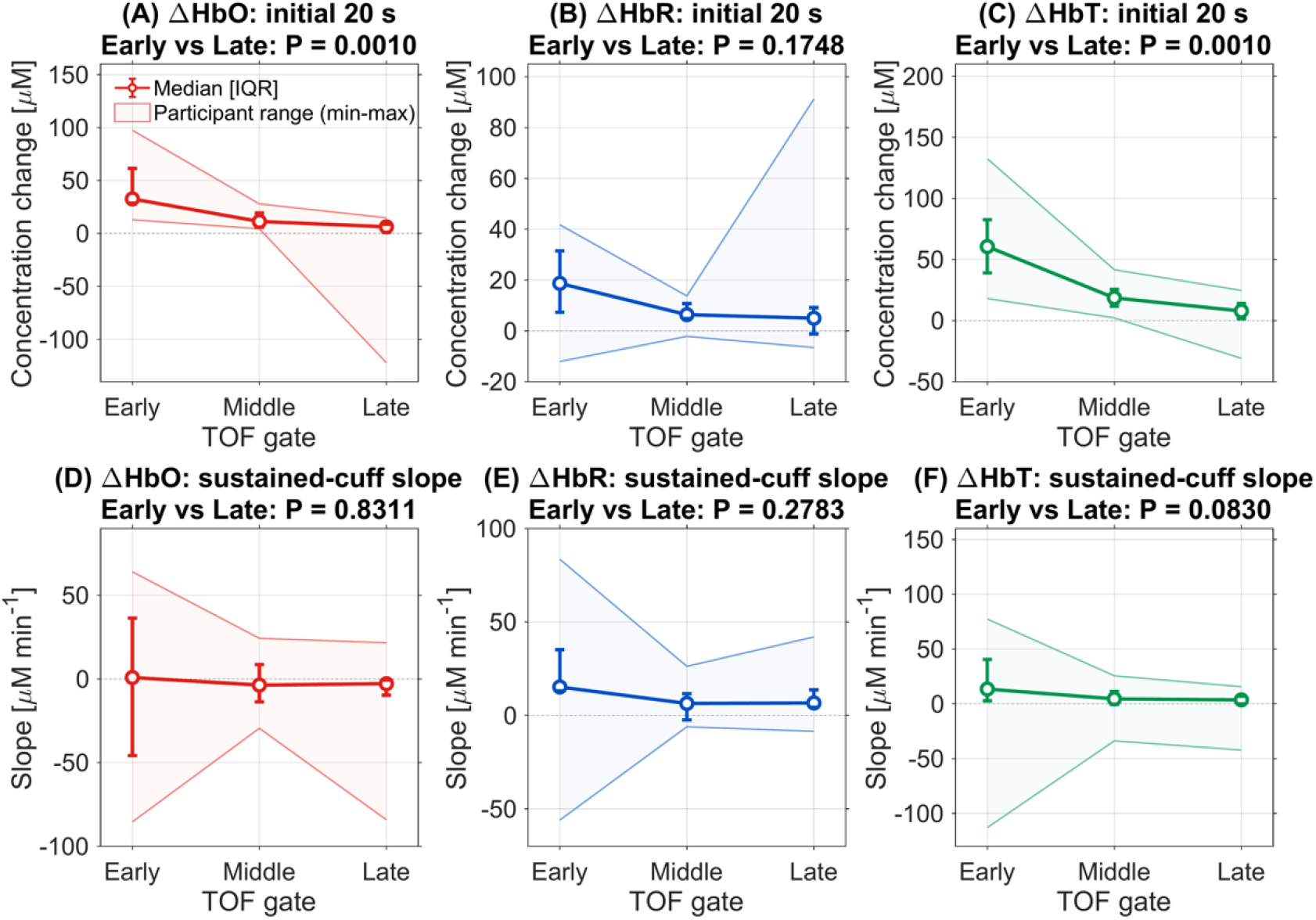
Phase-resolved hemoglobin responses across TOF gates (N = 11 independent participants). (A–C) ΔHbO, ΔHbR, and ΔHbT amplitudes during the first 20 s after cuff inflation, expressed in µM. (D–F) Sustained-cuff slopes of ΔHbO, ΔHbR, and ΔHbT, estimated by linear regression from 20 s after cuff inflation to 10 s before cuff release and expressed in µM min^−1^. Transparent color-matched envelopes and their boundary lines show the participant range (min–max). Colored lines, open circles, and error bars show median [IQR]. *P* values are from exact two-sided Wilcoxon signed-rank tests comparing the Early and Late TOF gates.

During the first 20 s, median ΔHbT was 63.9 µM in Early, 19.8 µM in Middle, and 5.8 µM in Late, corresponding to an approximately 11-fold Early-to-Late difference. Both the ΔHbT and ΔHbO Early-to-Late contrasts yielded P = 0.0010, whereas the corresponding ΔHbR contrast was not significant (P = 0.1748).

During sustained occlusion, the hemoglobin slopes were smaller and more heterogeneous. HbO tended to decrease and HbR to increase in the Late gate, but the paired Early-to-Late slope differences were not significant for HbO, HbR, or HbT. The longer-duration P01 control was consistent with a transition from an initial vascular-filling response toward the HbO-decrease/HbR-increase pattern expected during prolonged ischemia. This control is presented as mechanistic support and was not included in cohort-level inference.

### 4.5 Cuff responses reveal systematic TOF-gate dependence

Figure 10 summarizes the primary cuff-response metrics at the fixed Early, Middle, and Late TOF gates. Cuff flow suppression and reactive-hyperemia amplitude generally increased toward later gates, whereas the attenuation response decreased sharply from Early to Late. This opposing gate dependence was observed at both wavelengths.

**Figure 10.**
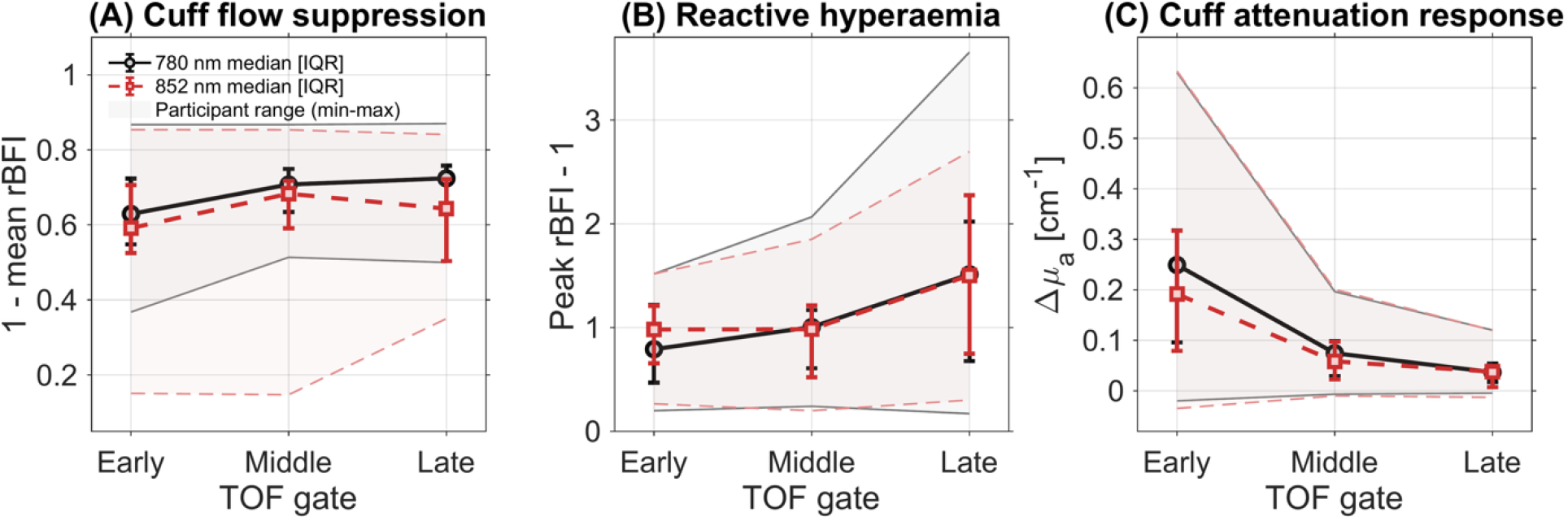
TOF-gate dependence of the primary cuff-occlusion responses (N = 11 independent participants). Participant-level metrics are summarized for the Early (75–125 ps), Middle (325–375 ps), and Late (525–575 ps) gates. (A) Cuff flow suppression, calculated as 1 minus the mean *κ*^2^-based rBFI during occlusion. (B) Reactive-hyperemia amplitude, defined as peak post-release rBFI minus 1. (C) Cuff-induced effective absorption change, Δ*µ*_*a*_. Symbols and error bars show median (IQR), while transparent envelopes and their boundary lines show the participant range (min–max). The 780 nm data are shown as black solid lines with circles and the 852 nm data as red dashed lines with squares.

The Late-minus-Early reactive-hyperemia contrast was +0.647 at 780 nm (95% CI 0.194 to 1.124; *P* = 0.0122) and +0.580 at 852 nm (95% CI 0.127 to 1.041; *P* = 0.0210). Conversely, the Early-minus-Late Δµ_*a*_ contrast was +0.196 cm^−1^ at 780 nm (95% CI 0.123 to 0.280) and +0.182 cm^−1^ at 852 nm (95% CI 0.100 to 0.274); both P = 0.0010. Cuff flow suppression also increased from Early to Late at 780 nm (+0.067; 95% CI 0.040 to 0.092; *P* = 0.0005), whereas the smaller 852 nm contrast did not reach significance (+0.036; 95% CI −0.034 to 0.095; *P* = 0.1030). Together, these metrics confirm that flow and attenuation responses are preferentially expressed in different photon-arrival-time windows.

## 5. Discussion

This study introduces dual-wavelength TOF-iSCOS as a unified interferometric approach for measuring wavelength-resolved attenuation, field dynamics, and photon time-of-flight. The attenuation and dynamic observables are derived from the same complex field and share the probe, detector, and acquisition clock, providing co-registration before physiological interpretation. The present 780/852 nm implementation also demonstrates how temporal multiplexing can extend the architecture to additional wavelength channels without duplicating the detection path.

The forearm cuff-occlusion measurements provide the principal *in vivo* validation. Flow suppression during occlusion and reactive hyperemia after release were recovered at both wavelengths. More importantly, the responses separated systematically across TOF gates: reactive-hyperemia rBFI increased toward the Late gate, whereas cuff-induced attenuation and the initial hemoglobin-volume response were strongest in the Early gate. Thus, TOF selection does more than alter signal strength or nominal sampling depth; it changes the relative weighting of distinct physiological processes. Flow dynamics, attenuation, and hemoglobin responses therefore provide complementary rather than redundant information.

The phase-resolved hemoglobin analysis further showed that the largest response amplitude did not necessarily identify the physiological phase of interest. Early-arriving photons emphasized the rapid vascular-filling response after cuff inflation, whereas later photons were more sensitive to the subsequent evolution of hemoglobin oxygenation. The longer-duration P01 control was consistent with this interpretation: Late-gate HbO transitioned from an initially positive response toward a slightly negative end-cuff response, while HbR remained elevated. Although this control provides mechanistic support rather than cohort-level validation, it demonstrates how occlusion duration and analysis window can alter the apparent hemoglobin response.

Direct access to the optical field also determines the estimator formulation. Interferometric detection measures *g*_1_ directly, while *κ*^2^*(T)* is reconstructed from the finite-integration visibility through an integral that is linear in *g*_1_. Both estimators recovered the cuff response, but the *κ*^2^-based approach retained substantially more usable fits in the photon-limited 852 nm Late gate. This improvement is consistent with the smoothing introduced by temporal integration and supports the use of *κ*^2^ as the primary estimator under the present photon budget. Because the two estimators use different lag ranges and exhibit rate-dependent differences, their fitted decorrelation rates should nevertheless be calibrated before being interpreted interchangeably.

The early-dominant attenuation response was observed at both wavelengths and was preserved in the TPSF-correction sensitivity analysis. The positive initial HbO and HbT responses are consistent with transient vascular filling after venous outflow is impeded, followed by increasing oxygen extraction during sustained occlusion. This sequence agrees with the reported dependence of the hemoglobin response on cuff duration [32] and with the longer-duration P01 control. The recovered hemoglobin amplitudes should be interpreted as effective, gate-resolved modified Beer–Lambert-law contrasts. Their magnitude depends on the gate-specific path-length estimate and the mixture of tissue layers contributing to each TOF window, rather than representing absolute bulk-tissue concentrations.

Temporal multiplexing provides wavelength co-registration but not strict simultaneity. The approximately 2.94 s wavelength-revisit interval was adequate for the relatively slow cuff response studied here. Faster functional paradigms will require shorter wavelength blocks, parallel detection, or an explicit assessment of interpolation and temporal-bandwidth effects. Late-gate performance also remains limited by photon availability. Increased collection efficiency, adaptive lag selection, and multi-channel averaging could extend the usable TOF range, with the expected trade-off between temporal resolution and signal-to-noise ratio.

The main limitations are the modest cohort size, single-channel geometry, relative rather than absolute flow scaling, and absence of an independent flow or oxygenation reference. The TOF gates remain instrument-convolved, and a single-exponential decorrelation model represents an effective description of mixed photon paths. Absolute TPSF-amplitude calibration and tissue-layer specificity were not established. Consequently, response magnitude should not be equated directly with muscle sensitivity. OEF and rOMI additionally depend on literature-assigned baselines and simplified compartmental assumptions and should therefore be regarded as exploratory relative indices. Future validation should combine improved photon collection with layered forward models, independent reference measurements, and larger cohorts.

Within these constraints, dual-wavelength TOF-iSCOS demonstrates that field decorrelation, spectral attenuation, and photon arrival time can be integrated within a single interferometric acquisition. The central finding is that flow, attenuation, and hemoglobin-phase responses can be preferentially expressed in different TOF gates. The latest or largest response is therefore not necessarily the most physiologically informative; wavelength, observable, TOF, and temporal phase must be considered jointly when interpreting diffuse optical measurements.

## 6. Conclusions

Dual-wavelength TOF-iSCOS provides co-registered measurements of flow-sensitive field dynamics and wavelength-resolved attenuation from a single interferometric acquisition. Across 11 independent participants, reactive hyperemia became more prominent in later TOF gates, whereas effective *Δµ*_*a*_ and the initial hemoglobin-volume response were weighted toward early-arriving photons. The *κ*^*2*^-based estimator improved fit retention under photon-limited Late-gate conditions, while phase-resolved hemoglobin analysis distinguished rapid vascular filling from the subsequent evolution of oxygenation.

These findings establish wavelength, field dynamics, photon arrival time, and physiological phase as complementary dimensions of diffuse optical contrast. Their joint measurement creates new opportunities to investigate how vascular filling, flow suppression and recovery, and oxygen extraction evolve across photon-path populations and temporal phases – processes that may be conflated in a single wavelength, TOF-averaged, or depth-averaged signal. TOF-iSCOS therefore provides a framework for designing measurements around the physiological process of interest rather than selecting a TOF window solely based on response magnitude.

The current dual-wavelength implementation can be extended to additional wavelength channels, opening a broader experimental space for studying coupled hemodynamic and metabolic responses in tissue.

## Data Availability

All data produced in the present study are available upon reasonable request to the authors.

## Acknowledgments

The authors thank InCellVu for providing in-kind support through access to laboratory space and the loan of computing hardware for data analysis and optical instrumentation for system calibration and characterization. InCellVu had no role in the study design, data analysis or interpretation, or preparation of the manuscript.

## Data and code availability

The analysis code and data are available upon reasonable request.

## Supplementary Information

### S1. Baseline effective optical properties

Baseline effective optical properties were estimated from TPSFs acquired during the 0.5–2.0-min baseline interval. After correction of frame-to-frame TPSF timing offsets, the baseline-averaged TPSF at each wavelength was fitted using a time-domain diffusion model for a homogeneous semi-infinite medium. The fits used the physical source-detector separation of 10 mm, a refractive index of 1.4, and wavelength-specific Gaussian instrument-response functions with measured FWHMs of 145.72 ps at 780 nm and 172.94 ps at 852 nm.

The diffusion-model fit yielded median [IQR] *µ*_*a*_ values of 0.078 [0.029–0.090] cm^−1^ at 780 nm and 0.101 [0.040–0.109] cm^−1^ at 852 nm, with corresponding *µ*_*s*_′, values of 11.69 [10.88–12.04] and 11.26 [9.78– 11.98] cm^−1^, respectively. Two participants reached the lower absorption-coefficient bound at both wavelengths; all finite fits were retained in the cohort summary. Because the forearm was represented by a homogeneous semi-infinite medium, these quantities should be interpreted as effective model parameters rather than layer- or compartment-specific optical properties.

**Figure S1.**
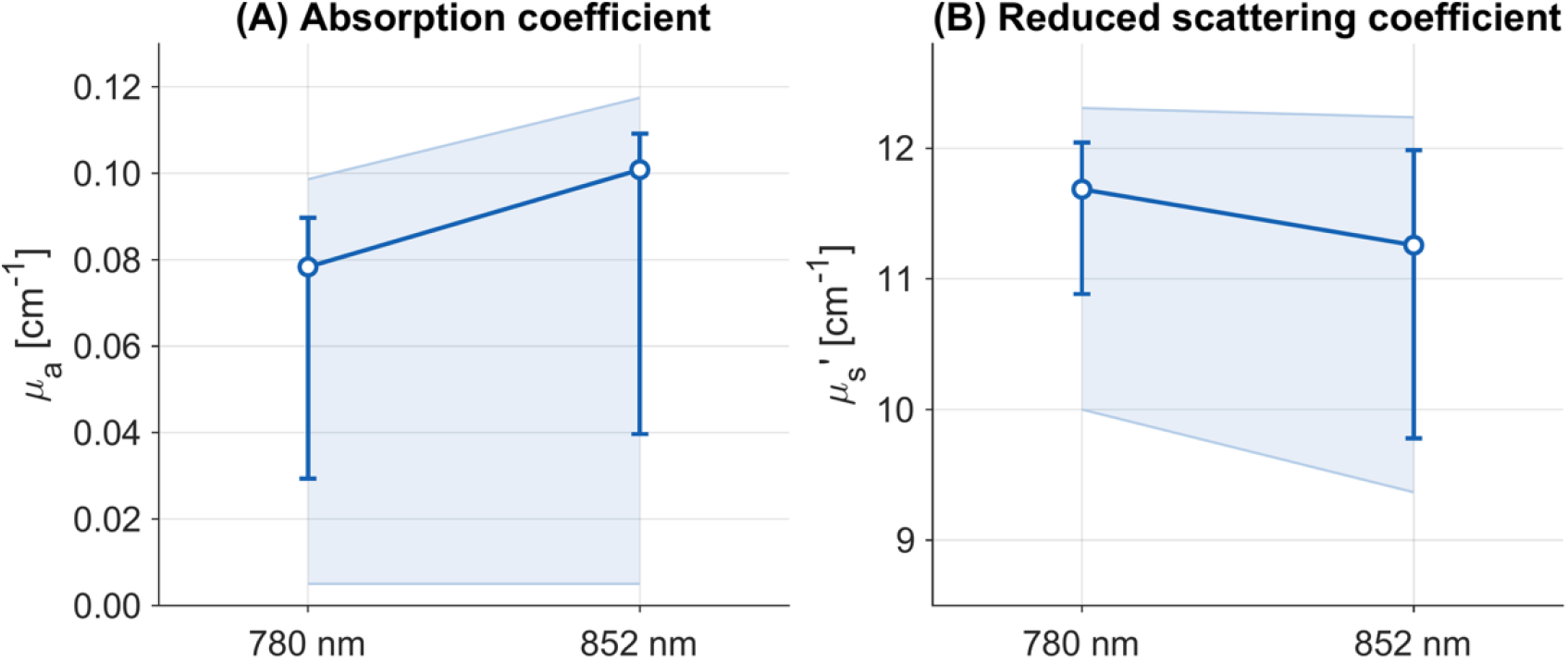
Baseline effective optical properties obtained by diffusion-model fitting (N = 11 independent participants). (A) Effective absorption coefficient, *µ*_*a*_, and (B) effective reduced scattering coefficient, *µ_s_* ′, at 780 and 852 nm. Baseline-averaged TPSFs were fitted using a semi-infinite time-domain diffusion model with the physical 10 mm source-detector separation and wavelength-specific measured instrument-response functions. Shaded regions and their boundary lines show the participant range (minimum–maximum); symbols and error bars show median [IQR]. Bilateral recordings from P01 were averaged before cohort summarization. All finite fits were retained.

### S2. Full-TPSF hemodynamic estimates

As a complementary analysis to the fixed-gate attenuation measurements, wavelength-resolved absorption changes were estimated from the shape of the complete TPSF. In this diffusion-model analysis, the baseline *µ*_*a*_ and *µ_s_*′, were first estimated for each participant and wavelength. Frame-wise *µ*_*a*_ was then fitted with *µ_s_* ′ fixed at its baseline value to reduce the covariance between the two optical properties.

**Figure S2.**
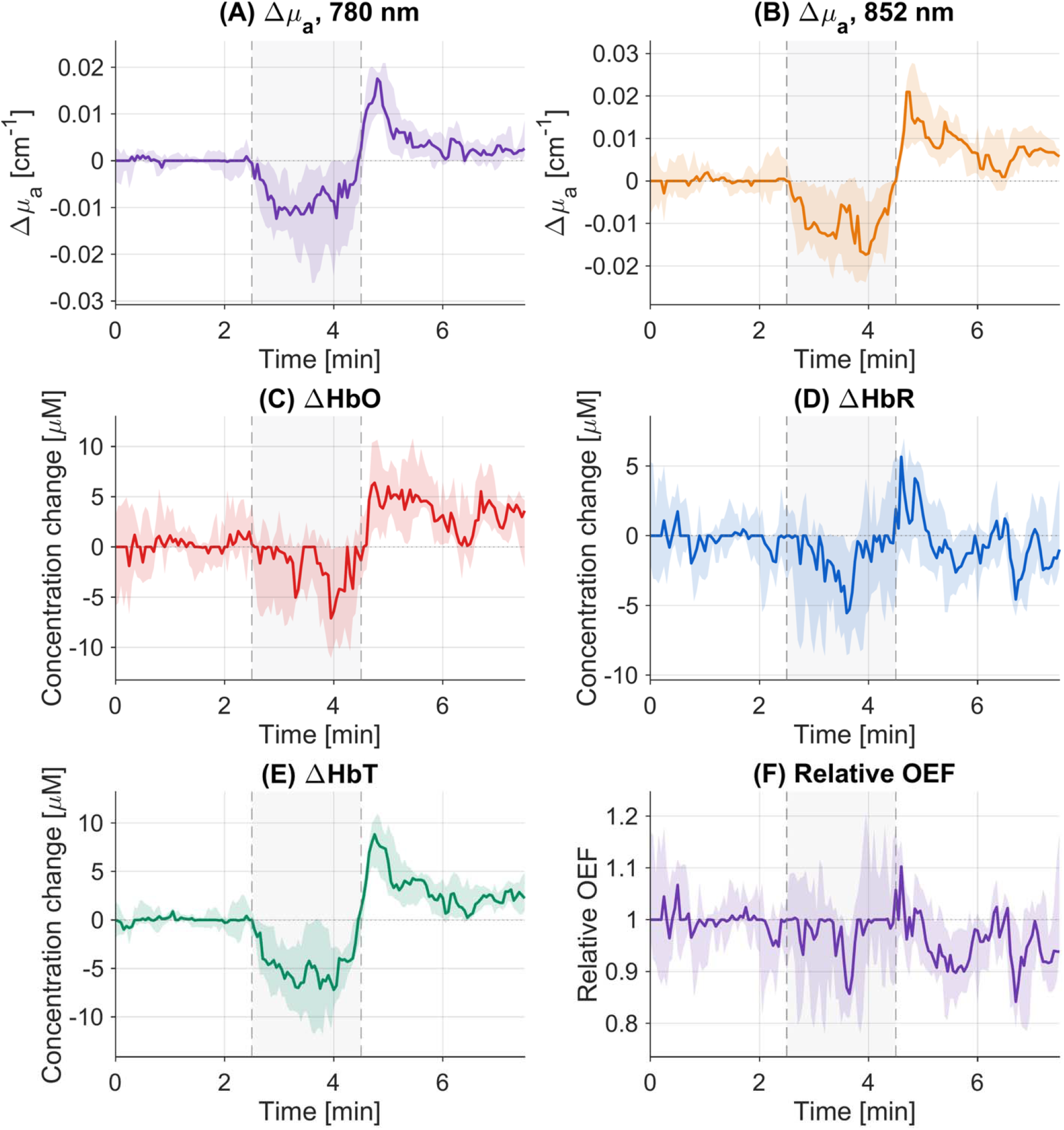
Full-TPSF hemodynamic trajectories obtained by diffusion-model fitting. (A,B) Changes in the effective absorption coefficient, *µ*_*a*_, at 780 and 852 nm relative to the 0.5–2.0 min baseline. For each frame, *µ*_*a*_ was fitted to the peak-aligned TPSF using the semi-infinite time-domain diffusion model, while *µ* was fixed to the participant- and wavelength-specific baseline estimate. (C–E) Corresponding ΔHbO, ΔHbR and ΔHbT obtained by two-wavelength inversion. (F) Oxygen-extraction fraction relative to baseline. Curves and color-matched shades show the participant median and IQR, respectively. The gray region indicates cuff inflation from 2.5 to 4.5 min; dashed lines mark inflation and release. Traces were smoothed using a seven-frame moving median. Bilateral recordings from P01 were averaged before cohort summarization.

The resulting Δ*µ*_*a*_ traces were converted to ΔHbO and ΔHbR using the two-wavelength extinction matrix, with ΔHbT calculated as their sum. Relative OEF was derived using the same baseline hemoglobin assumptions as in the main analysis. The diffusion-model estimates showed a decrease in effective *µ*_*a*_ during cuff inflation followed by a pronounced post-release rebound at both wavelengths. The hemoglobin inversion amplified differences between the wavelength channels, particularly for HbO and HbR, whereas HbT retained the more consistent cuff-related decrease (Fig. S2).

These results should be interpreted as full-TPSF, model-dependent observables rather than gate-resolved tissue concentrations. Their behavior differs from the intensity-derived fixed-gate attenuation response because the optical-property approaches are driven primarily by changes in TPSF shape, whereas the gate analysis also retains changes in detected amplitude.

Although the optical-property analysis used a homogeneous semi-infinite model, the resulting hemoglobin trajectories retained physiologically plausible temporal structure. In particular, the diffusion-fit estimates showed a cuff-related decrease in ΔHbT followed by a rapid post-release increase, consistent with the suppression and subsequent restoration of blood volume within the sampled tissue. ΔHbO exhibited a comparable post-release overshoot, as expected from the influx of oxygenated arterial blood during reactive hyperemia. These relative changes are likely more robust than the absolute fitted optical properties because participant-specific, wavelength-dependent modeling biases are largely removed by baseline subtraction. Nevertheless, the homogeneous model cannot distinguish contributions from skin, adipose tissue, muscle and superficial vessels, and changes in the relative sampling of these compartments may be absorbed into the fitted *µ*_*a*_. The agreement of the principal ΔHbT and ΔHbO features with the expected cuff-response sequence therefore supports their physiological relevance, while their absolute amplitudes should be interpreted as effective, model-dependent estimates rather than compartment-specific concentration changes.

### S3. Rate-dependent estimator comparison

Across the accepted stage-, wavelength- and gate-resolved pairs, the reconstructed-*κ*^2^ and direct-*g*_1_ decorrelation rates were strongly correlated (*r* = 0.9939). The relationship was nevertheless not one of strict estimator identity. The log–log slope of 1.111 and the quartile analysis in Fig. S3B indicate a systematic rate dependence: reconstructed-*κ*^2^ estimates were generally lower at slow decorrelation rates, particularly during cuff occlusion, and approached the direct-*g*_1_ estimates as the rate increased.

Baseline normalization substantially reduced this calibration difference, and both estimators preserved the principal physiological sequence of flow suppression during cuff inflation and reactive hyperemia after release (Fig. S3D). The reconstructed-*κ*^2^ route also retained a higher accepted-frame fraction in photon-limited conditions, with the clearest difference occurring in the 852 nm Late gate. These acceptance fractions were calculated using each estimator’s predefined fitting interval and should therefore be interpreted as pipeline-level yield rather than a controlled comparison of their intrinsic statistical efficiency.

**Figure S3.**
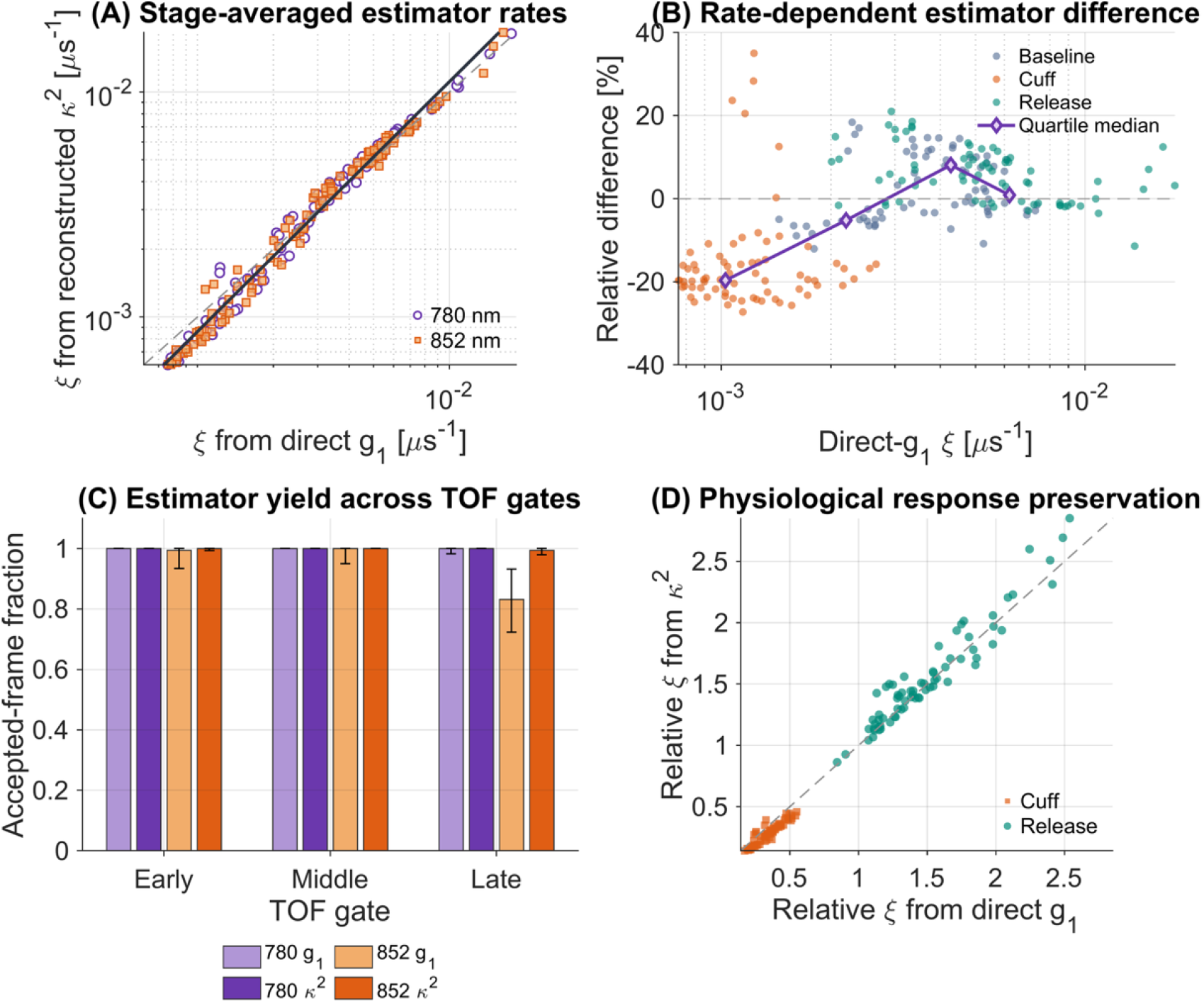
Rate-dependent comparison of the direct-*g* and reconstructed-*κ*^2^ decorrelation-rate estimators. (A) Stage-averaged *ξ* estimates for accepted wavelength-, gate- and protocol-stage pairs. Data at 780 nm are shown as violet circles and data at 852 nm as orange squares. The dashed line denotes identity and the solid line shows the log–log regression. (B) Relative estimator difference as a function of the direct-*g*_1_ rate. Colors identify baseline, cuff-occlusion and release stages; the violet line connects the median differences within quartiles of the direct-*g*_1_ rate. (C) Participant-level accepted-frame fractions across TOF gates. Bars and error bars show median [IQR] across 11 independent participants after averaging the bilateral P01 recordings. Lighter bars represent direct-*g*_1_ fitting and saturated bars reconstructed-*κ*^2^ fitting. (D) Cuff and post-release responses after normalization to the corresponding baseline rate. The dashed line denotes identity.

Part of the remaining estimator difference may arise from the distinct lag domains used for fitting. The direct-*g*_1_ and reconstructed-*κ*^2^ estimators weight different portions of the decorrelation curve, and their fitted rates therefore need not be numerically identical even when they describe the same underlying field dynamics. Agreement could potentially be improved by jointly selecting the fitting windows to provide more closely matched sensitivity to decorrelation rate or by applying an empirical cross-estimator calibration. Such optimization would, however, involve a trade-off between numerical agreement, fit stability and accepted-frame yield, particularly in photon-limited Late gates. We therefore retained the independently selected and prospectively frozen fitting domains in the present analysis rather than tuning them to maximize agreement in the study cohort. The observed rate-dependent relationship provides a basis for developing such calibration in future measurements.

### S4. Reconstructed-*κ*^2^ fit-window sensitivity

The reconstructed-*κ*^2^ fitting domain was selected using a fine-sampling 780-nm development recording before analysis of the forearm cohort. Candidate windows varied both the initial lag and the end of the fitting interval. The prespecified quality score jointly considered fit quality, baseline stability and accepted-frame yield, while the Late-to-Early hyperemia ratio was evaluated separately to confirm preservation of the expected TOF-dependent physiological response (Fig. S4).

**Figure S4.**
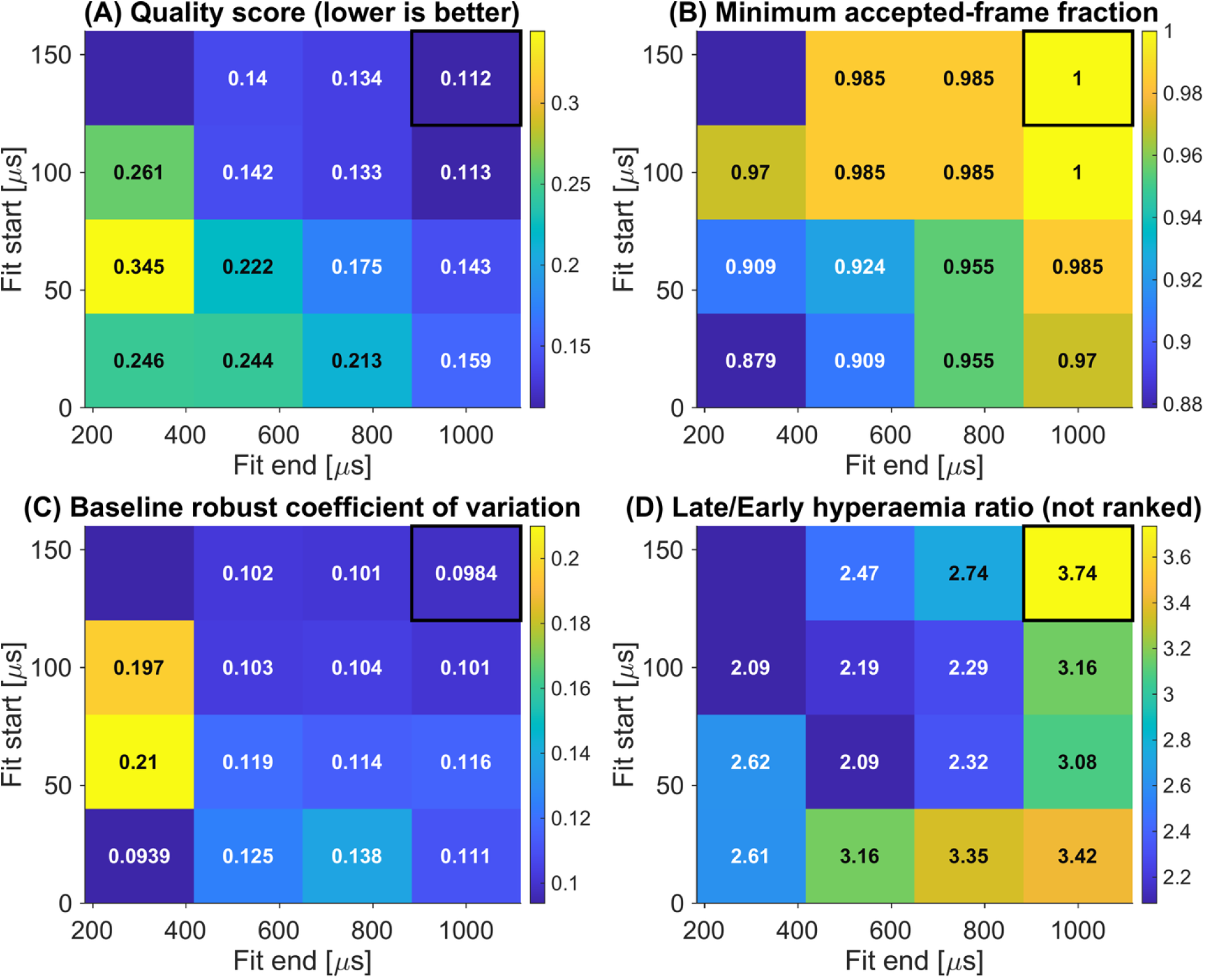
Reconstructed-*κ*^2^ fit-window sensitivity in the 780-nm development recording. Candidate fitting domains were defined by the fit-start and fit-end lags. Heat maps show (A) the prespecified composite quality score, for which lower values indicate better overall performance; (B) the minimum accepted-frame fraction across the analyzed TOF gates; (C) the median baseline robust coefficient of variation; and (D) the Late-to-Early reactive-hyperemia ratio, which was retained as a physiological-response check but was not included in the numerical ranking. Values are reported within individual cells. The black rectangle identifies the selected 140-1000-µs fitting domain, which was frozen before cohort analysis.

The selected 140-1000 µs domain achieved the lowest composite quality score (0.112), a minimum accepted-frame fraction of 1.000 and a baseline robust coefficient of variation of 0.0984, while retaining a Late-to-Early hyperemia ratio of 3.74. This relatively late and extended fitting interval avoids the short-lag region most affected by finite-integration reconstruction while retaining sufficient dynamic range for stable rate estimation. The domain was subsequently frozen and applied without participant-specific adjustment.

The direct-*g*_1_ fitting window was selected using the same development-data framework, but its largely analogous sweep is not reproduced to avoid duplicating methodological detail. The resulting 20-700 µs domain is reported in the Methods, and its cohort-level behavior is compared directly with the reconstructed-*κ*^2^ estimator in Fig. S3.

